# Programmatic implementation of kangaroo mother care: a systematic synthesis of grey literature

**DOI:** 10.1101/2023.04.05.23288153

**Authors:** Anne-Marie Bergh, Shuchita Gupta, Suman Rao

## Abstract

**Background:** Evidence on the effectiveness of kangaroo mother care (KMC) is available and guidelines have been formulated. However, little is known about the programmatic implementation of KMC at national and subnational levels.

**Methods:** A structured search of grey literature was conducted to identify reports of national or subnational level implementation of KMC to understand the population-based coverage of KMC, availability of KMC services at national and subnational levels, programmatic approaches to scale, and health systems actions that may influence KMC scale-up. The search strategy included two rounds of screening using a variety of grey literature search engines, portals, repositories, and targeted websites, as well as snowball sampling. Data from 212 documents were extracted and transferred into a database with an extensive topic list. These documents were then classified as “for in-depth analysis”, “possible nuggets”, and “not important”. Further analysis of 42 information-rich documents was conducted with NVivo software to identify recurring themes.

**Findings:** We found information on KMC implementation on a national or subnational scale for 18 countries. Estimates for national- or subnational-level population-based coverage of KMC were available from only six countries (Colombia 63%, the Philippines 53%, Malawi 22%, Bangladesh 22%, India 13%, Viet Nam 8%). Information on the availability of KMC services was scant and fragmented, with no information on their quality. Programmatic KMC implementation is characterised by leadership from a variety of implementation partners and by different implementation trajectories. Countries for which information on KMC implementation was available shared common health system actions such as the inclusion of KMC in national policy, recognition of KMC as a priority newborn health intervention and strong advocacy by champions at all levels, dedicated investment in KMC and in some cases insurance cover for KMC, capacity building and motivation among health workers, dedicated space for KMC with equipment and supplies, support for KMC practice, and data collection and use.

**Conclusion:** Programmatic implementation of KMC requires action in multiple health system building blocks with a focus on monitoring and evaluation of availability and quality of services, along with coverage.

---

Kangaroo Mother Care (KMC) is a method of care for preterm and low birthweight (LBW) infants that reduces neonatal morbidity and mortality [1]. The World Health Organization (WHO) defines KMC as continuous and prolonged skin-to-skin contact of the baby with a caregiver and support for exclusive breastmilk feeding [2]. Two other features that are widely considered to be part of the KMC package are early discharge from hospital with a reliable follow-up system in place and support by healthcare workers, the family and the community [1, 3–5].

KMC is a complex intervention [6] that depends on strengthening of health systems and on human behaviour [5, 7]. In addition, facility-based KMC forms an integral part of the care packages for small and sick newborns [8]. Where KMC was originally largely seen as an add-on intervention that was introduced vertically in a small number of higher-level health facilities [9], it gradually became more embedded in the wider focus of strengthening the care of small and sick newborns and promoting family-participatory care, especially at the level of district hospitals and other lower-level health facilities [8, 10–13].

Although there are many claims and reports about the successful implementation of facility-based KMC, little is known about the programmatic implementation of KMC and the availability of KMC services globally or at national or subnational levels. Data in the published domain provide insufficient information on population-based coverage of KMC. As KMC is not yet being captured in the health management information system (HMIS) of most countries, some form of review of the grey literature could help identify countries with some footprint of programmatic KMC implementation and reveal how health system and process factors have affected their implementation and scale-up activities. The objective of this review was to understand and synthesise information on population-based coverage of KMC, the availability of KMC services at national and subnational levels, programmatic approaches to scale, and health system components and process factors that may influence scale-up of KMC.

## METHODS

The initial search question was framed very broadly: How is programmatic implementation of KMC reported in the grey literature? The structured search also endeavoured to identify packages of health interventions that had achieved high coverage of population-based KMC (>50%) as well as details of the components of such packages.

Coverage is defined as the proportion of people (preterm or LBW babies) who received a specific intervention or service (KMC) in a particular geographic or administrative region [14]. Implementation is interpreted as the extent to which a programme is delivered as intended – there are individual-level (mother–infant dyad) and programme-level (KMC services) measures of implementation [15]. Scale-up is defined as the programmatic expansion of KMC services to bring the benefits of KMC to more preterm and LBW babies over a wider geographical area more quickly and more equitably, over longer periods [16, 17].

### Eligibility criteria

Finding documents in the grey literature on the programmatic implementation of KMC involved broaching uncharted territory. The initial search net was cast very wide before being narrowed down. Grey literature was interpreted as including any of the following types of documents: documents related to conferences and meetings (e.g. abstracts, presentations, posters, summaries, working group reports); government documents (e.g. policy statements, guidelines, strategic plans); project reports, reviews and briefs; assessments and evaluations; newsletters; blogs; miscellaneous slide presentations; and unpublished dissertations or theses.

The setting of inclusion and exclusion criteria was guided by what could realistically be managed in one review. Only printable electronic documents and documents with “kangaroo care” or “kangaroo mother care” in the title or abstract were included (French: “mère kangourou” or “maman kangourou”; Spanish: “madre canguro”; Portuguese: “mãe canguru”). Video and audio recordings and terms related to the individual components of KMC (skin-to-skin, breastfeeding, support, discharge and follow-up) and the care of small and sick newborns were excluded.

As the purpose of the review was not to find or grade evidence on the effectiveness of interventions to implement or scale up KMC programmes, criteria such as risk of bias and certainty assessments were not applicable.

### Information sources

Grey literature was found in two ways: through internet searches and through snowball sampling by contacting individuals working in the field. Figure 1 provides an overview of the screening and data analysis processes that took place between June and December 2021.

**Figure 1.**
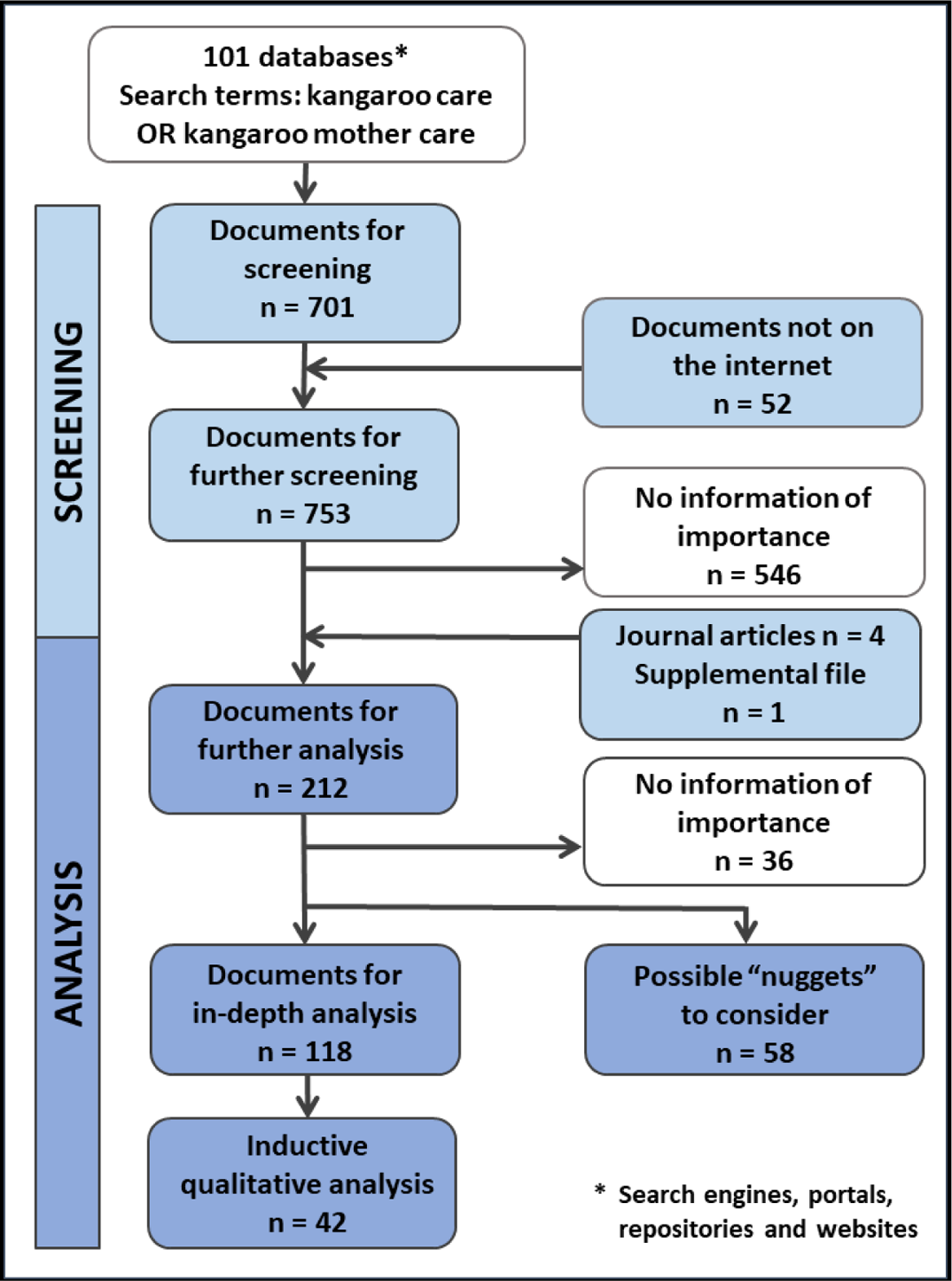
Flow diagram of the search and analysis strategies.

### Search strategy

The search strategy consisted of two phases of screening. Initially, a very broad exploratory search was conducted on the internet using search engines, portals, repositories and websites (henceforth called “databases”) until no new links to databases were found. The primary screening route is described in supplemental file 1. The databases included OpenDOAR and university websites with extensive links to other grey literature databases (Illinois Chicago Public Health; Monash; La Trobe). In addition to these searches, additional sources were specifically targeted for searching, such as the Kangaroo Care Bibliography which is regularly updated by Susan Ludington and the following websites: *Fundación Canguru* in Colombia which manages the documents of the International Network of Kangaroo Mother Care (INK); UNICEF; development agencies known for their involvement with kangaroo mother care implementation (e.g. Save the Children, USAID, jhpiego, JSI Research & Training Institute, Inc.); and specific programmes ( e.g. Saving Newborn Lives, Every Preemie-Scale, ACCESS, MCHIP, MCSP). Search histories with potentially relevant titles were saved as pdf files. The internet search was conducted on 101 databases (supplemental file 2) and 701 documents were identified for a second round of screening. The number of hits per database ranged from 0 to 8185.

During the first screening, another 52 documents that are not available in the open domain on the internet were added: 42 documents received from individuals and 10 documents the reviewer had at her disposal.

Of the 753 documents identified for a second screening, 499 documents were retrieved from five of the specifically targeted websites, with 323 from the INK website. When the 753 documents’ tables of contents and their “Find” functions were used, a second screening entailed a manual search for terms related to “coverage”, “implementation” and “scale-up”, which yielded 212 documents. These documents reported on research or project outcomes, other kinds of assessments or any means of describing implementation processes or experiences.

### Data collection, management and extraction

For easier management, the 212 remaining documents were grouped as follows: (AB) Conference abstracts, posters, summaries and working groups (n=49); (AS) Assessments, evaluations and reviews (n=31); (R) Reports, briefs, newsletters and blogs (n=53); (SP) Slides and conference presentations (n=60); and (SC) KMC implementation and scale-up documents (n=19). A database was created in MS Excel to summarise the relevant information gleaned from the documents and identify the documents likely to be of greater interest. Supplemental file 3 contains a summary of the process followed to complete each column in the database. Regarding KMC coverage and components of KMC implementation, a classification of A-E was included to get a sense of the distribution of these important concepts in the documents.

The reviewed documents were marked in the database as follows: (1) red for further in-depth analysis (n=118); (2) yellow for possible “nuggets” to consider (n=58); and (3) white for nothing of importance (n=36). The documents marked with red and yellow were further reviewed to identify documents that were sufficiently information-rich to be included in an inductive qualitative analysis using NVivo9 software [18]. Thirty-seven (37) grey documents were uploaded onto NVivo, together with four published articles and one article supplement that had been brought to our attention during the review (supplemental file 4). We identified 70 categories (nodes) in this analysis (see supplemental file 5) and they were used to identify emerging factors that played a role in country implementation of KMC. Supplemental file 6 contains a further breakdown of the major document types identified in the analysis.

### Synthesis of findings

The approach to the synthesis included extracting numbers, but also an analysis along the lines described by Spicer et al describe in their article, “Scaling-up is a craft not a science” [19]. Results were synthesised around two main themes: the numbers and the manner (the craft). Numbers are given with regard to population-based coverage of KMC and the availability of KMC services on a national or subnational level (district, region, province, state). For the numbers synthesis, we included published results that were not part of the grey literature review.

The manner in which countries approach KMC implementation and scale-up includes the “what” and the “how”: health system factors (building blocks) and contextual factors (people as catalysts for implementation and process as a pathway to implementation). These factors could also act as enablers of or barriers to successful implementation. Figure 2 is a snapshot of the interrelatedness of factors involved in the programmatic implementation of KMC.

**Figure 2.**
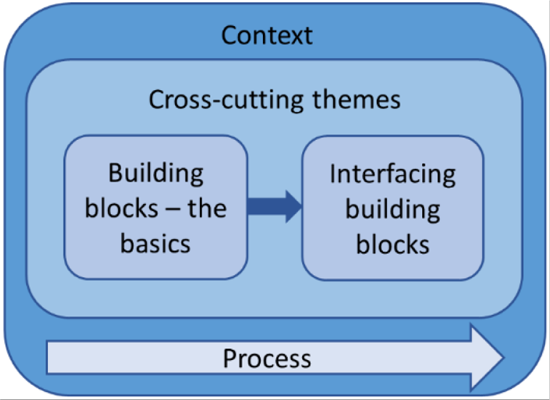
The interrelatedness of factors involved in KMC implementation at scale.

## FINDINGS

### Programmatic implementation of KMC and availability of KMC services at a national or subnational level

Until around 2012, there was little movement with regard to implementing KMC on a large scale in most countries – in the grey literature, information on early scale-up was only available from Brazil [20–22], Malawi [23, 24], and South Africa [25, 26]. In 2018, 28 (32%) of the 90 countries that provided data for the Every Newborn Action Plan (ENAP) database reported having an updated national policy or guideline on KMC [27]. Project reports, conference presentations and publications suggesting that a country had implemented KMC at a national or subnational level were available from 17 ENAP countries: Bangladesh, Brazil, Ethiopia, Ghana, India, Indonesia, Iran (Islamic Republic), Malawi, Mali, Nepal, Nigeria, the Philippines, Rwanda, South Africa, Tanzania, Uganda and Viet Nam. In addition, information on KMC implementation was also available for Colombia. KMC implementation on a subnational scale was defined as implementation of KMC in at least a group of hospitals and health centres, not necessarily located in the same geographic or administrative area.

Information on KMC service availability was scarce and fragmented. Most reports did not refer to continuation of services or sustainability beyond the point of assessment or measurement. From the available documents at the time of the review we found that KMC services were reported to be available as follows: in the majority of departments of Colombia (2021) [28]; across 43% of national, regional, and provincial hospitals in the Philippines (2019) [29]; across 108 subdistrict health complexes and higher-level facilities (district hospitals and medical college hospitals) in urban and rural areas in Bangladesh (2020) [30]; in 11 districts in Malawi (2017) [31]; in 18 of the 63 provincial and three national hospitals in Viet Nam (2015) [32]; in more than 80% of the hospitals in the North West Province of South Africa (2015) [33]; in 53% of level-3 hospitals in the Islamic Republic of Iran (2012) [34]; and in 121 health facilities in Malawi (2012) [24].

We could only find one grey literature document reporting KMC scale-up in high-income countries. A cross-national survey performed in around 2018 among level-III neonatal units in Spain reported that 71% of the responding neonatal intensive care units practised kangaroo care [35].

Despite information on availability of KMC services, there were questions about the uptake and quality of services [33, 36]. In addition, there did not seem to be consensus on the minimum requirements for a KMC service at different levels of care.

### Population-based coverage of kangaroo mother care: reliable estimates lacking in most countries

In 2017, 20% of the 90 countries contributing to the ENAP database reported having an indicator in their national HMIS for the number of babies <2000 g initiated on KMC in facilities with KMC services [37]. However, KMC coverage estimates for preterm or LBW newborns (<2000 g or <2500 g) are lacking in most countries, even in countries that have implemented KMC on a national or subnational scale.

The review of the 212 documents found three documents we had classified as A (quantitative information on high population-based KMC coverage, >50%), reporting population-based coverage of KMC from two countries (Colombia and the Philippines) and one district in Malawi. For *Colombia*, data on KMC coverage was available for the period 2015–2021. In 2015 it was estimated that in four cities more than 50% of eligible newborns had access to KMC (implying that they had received continuous KMC), with the estimates for five more cities ranging between 22% and 44% [38]. In 2018 it was estimated that five cities had a coverage of more than 80%; another 19 had a 50–80% coverage; and three had under 50% [39]. An updated document received in 2022 estimates that 63.2% of eligible babies had access to KMC in 2021 [28].

By 2019 an estimated 53% of preterm and LBW babies were receiving KMC in the *Philippines* [29]. Only one district in *Malawi* (the Thyolo district) reported a coverage of more than 50% in 2016 (64%), and this dropped to 49% in 2017 [31]. The national KMC coverage (measured as the number of expected cases of babies <2000 g initiated on any KMC) was estimated to be 22% in 2017, based on 2014–2017 data from 11 of 28 districts [31, 40]. However, data extracted from maternity reports onto the District Health Information Software 2 (DHIS2) in 2021 suggest that fewer than 10% of preterm or LBW newborns in Malawi received facility-based KMC [41].

With regard to documents classified as B (quantitative information on ANY KMC coverage), four documents on facility-based populations were found in the grey literature for the period 2015–2017. A *Rwanda* presentation reported facility-level KMC admission rates of LBW newborns in 10 districts in the 2016 financial year as being 55%, 68%, 61% and 46%, for the four quarters respectively [42]. From 1 January 2015 to 31 July 2017, about 800,000 newborns were treated in special newborn care units in *India*, of whom 50,959 (13.2%) received KMC, with variations from 0–31% between states, according to a report for a working group [43]. A conference abstract from *Viet Nam* indicated that in 2015, according to routine national data, 8.2% of LBW newborns received KMC; however data from seven provincial hospitals showed a 22% rate [32]. In two KMC district-hospital demonstration sites established through the *Vriddhi* project in *India,* an increase in KMC initiation from 25% to 45% to 63% was reported over three quarters of 2017 [44].

According to a report on data use in Malawi, which has KMC indicators in its DHIS2, “data completeness and timeliness remain a challenge” (p1) [40]. A brief on a multi-country evaluation in Africa referred to the importance of achieving an equilibrium so that services are not expanded too rapidly at the expense of quality care [36].

### Health system factors involved in the programmatic implementation of KMC at scale: what countries have done

Because of the lack of information available on high-coverage, population-based KMC in the grey literature and on what these packages entailed, we used the health systems building blocks to organise the key factors reported to have facilitated large-scale implementation of KMC in seven countries for which sufficient information was available. Our findings are presented in Table 1. The categorisations in two published papers were combined into seven building blocks to organise the findings [11, 45]. It should be noted that in the same country, subnational entities may have different levels of implementation success, depending on the level of prioritisation of the care of small and sick newborns and various contextual, geographic, social, economic, and political factors outside the health system. The seven countries referred to are at different stages of national scale-up and some of the lessons listed in the table were learned within projects that had not been rolled out nationally at the time of reporting. If a country is not included in the table or an included country does not have a grey dot for a particular lesson, it merely means that information was not found in the documents consulted for the grey literature review.

**Table 1.** Lessons from national KMC programmes by health system building block

| HEALTH SYSTEM<br>BUILDING BLOCK | FACTORS THAT FACILITATE KMC IMPLEMENTATION | COUNTRY |  |  |  |  |  |  |
| --- | --- | --- | --- | --- | --- | --- | --- | --- |
|  |  | Ban | Bra | Col | Ind | Mal | Phil | SA |
| Policy, governance<br>and leadership | Adoption of KMC in national or subnational policy and guidelines | ● | ● | ● | ● | ● | ● | ● |
|  | Advocacy by champions (e.g. professional organisations and political leaders) at national and subnational levels and linkage with international KMC networks | ● |  | ● | ● | ● | ● | ● |
|  | Recognition of the value of KMC as a priority newborn health intervention | ● |  | ● | ● | ● |  |  |
| Health system<br>financing | Dedicated funding/investments for KMC from government and/or donors | ● | ● | ● | ● | ● | ● | ● |
|  | Inclusion of KMC as an essential maternal and newborn health service covered under insurance (partially or fully) | ● | ● | ● | ● |  | ● |  |
| Health workforce | Capacity-building of health workforce, including pre-service and in-service training and sensitisation on KMC | ● | ● | ● | ● | ● | ● | ● |
|  | Motivation of staff and creation of a network of health care professionals | ● |  | ● | ● |  | ● |  |
|  | Facility staff serving as champions and motivators | ● |  | ● |  |  |  | ● |

| HEALTH SYSTEM BUILDING BLOCK | FACTORS THAT FACILITATE KMC IMPLEMENTATION | COUNTRY |  |  |  |  |  |  |
| --- | --- | --- | --- | --- | --- | --- | --- | --- |
|  |  | Ban | Bra | Col | Ind | Mal | Phil | SA |
| Health service delivery | Provision of family-centred care |  | ● | ● | ● | ● | ● | ● |
|  | Clarity about KMC and its practice (e.g. protocols, counselling board, tools, job-aids) at each level of care | ● |  | ● | ● | ● |  | ● |
|  | Centres of excellence and/or champions for KMC in tertiary and secondary level facilities (i.e. coaches to train others) | ● |  | ● | ● |  | ● | ● |
|  | Continuity of care within the health system and between the facility and community (including referral system) | ● |  | ● |  |  |  | ● |
|  | Post-discharge follow-up at a health facility or in the community | ● |  | ● | ● |  | ● | ● |
|  | Integration of KMC into SSNC, comprehensive newborn care and/or maternal and newborn care packages |  |  | ● | ● | ● | ● | ● |
|  | Utilisation of service readiness surveys or situational analyses to inform the planning of scale-up | ● |  |  |  | ● |  |  |
|  | Target setting for KMC coverage | ● |  |  |  |  |  |  |
| Health infrastructure and supplies | Dedicated space for KMC with equipment and supplies | ● |  | ● | ● | ● | ● | ● |

| HEALTH SYSTEM BUILDING BLOCK | FACTORS THAT FACILITATE KMC IMPLEMENTATION | COUNTRY |  |  |  |  |  |  |
| --- | --- | --- | --- | --- | --- | --- | --- | --- |
|  |  | Ban | Bra | Col | Ind | Mal | Phil | SA |
| Health management information systems | Monitoring KMC through HMIS using specific indicators | ● |  |  | ● | ● |  | ● |
|  | Creation of a data collection platform (not at national level) with specific indicators for access and outcomes |  |  | ● |  |  | ● |  |
|  | Introduction of data management systems for data use and sharing to ensure data quality and utilisation of data for action | ● |  | ● | ● |  |  |  |
|  | Capacity-building on data collection, analysis, and interpretation | ● |  | ● |  |  |  |  |
| Community ownership and partnership | Social and behaviour change communication initiatives and community awareness building | ● |  |  | ● | ● | ● |  |
|  | Use and orientation of existing community health workers and support groups for referral and post-discharge follow-up | ● |  |  | ● |  |  |  |
|  | Engagement of community structures and community health workers for community-based provision of KMC |  |  |  | ● |  |  |  |

**COUNTRIES:** Bang, Bangladesh; Bra, Brazil; Col, Colombia; Ind, India; Mal, Malawi; Phil, the Philippines; SA, South Africa.
**Abbreviations:** HMIS, health management information system; KMC, kangaroo mother care; SSNC, small and sick newborn care.
Found in at least one document
No information found

### Contextual factors influencing the programmatic implementation of KMC at scale: how countries have done it

Contextual factors cut across health system building blocks. Two main factors emerged: people as catalysts in implementation and process as pathways to implementation. People are key in driving programmatic KMC implementation and there is a need for appropriate capacity development. Then there are processes of KMC implementation that determine different pathways at different levels of national and subnational health systems and across countries.

#### People: catalysts in driving programmatic KMC implementation

Enthusiastic, committed individuals or passionate drivers (individuals, groups, agencies) are mentioned as the key to implementation success, be it at facility level, subnational or national level or at a broader regional or global level [46–51]. People play a role in leadership and governance, health system financing, the health workforce, health service delivery, and community ownership and partnership.

Various terms are used interchangeably in the grey literature to refer to individuals or groups of people as catalysts or change agents: stakeholders; role players; drivers; champions [16, 52–55]. We synthesised these under the umbrella term of “implementing partners” in Table 2, each with a number of topics to describe possible influences. The topics included people, entities, resources, support, and challenges.

**Table 2.**
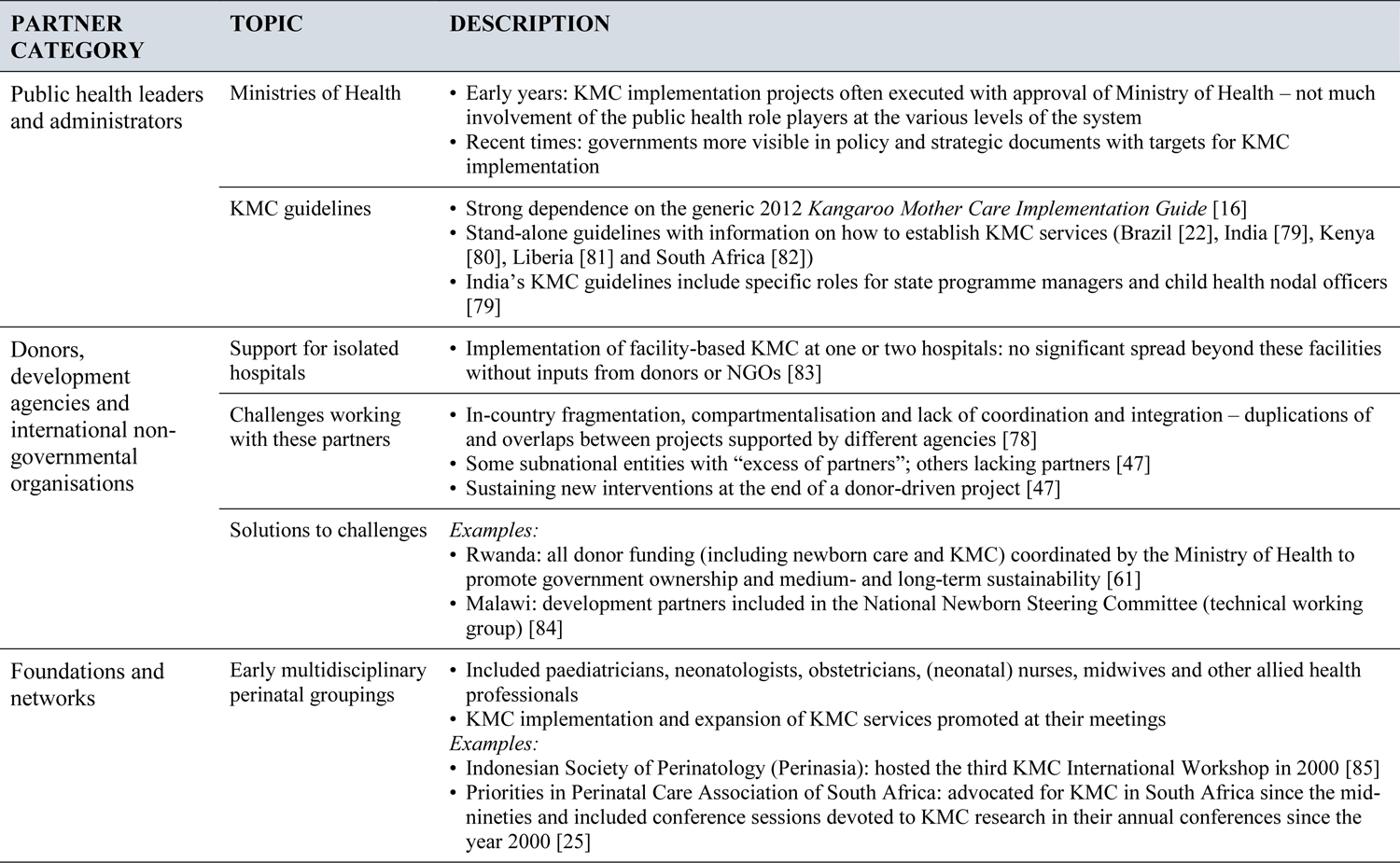
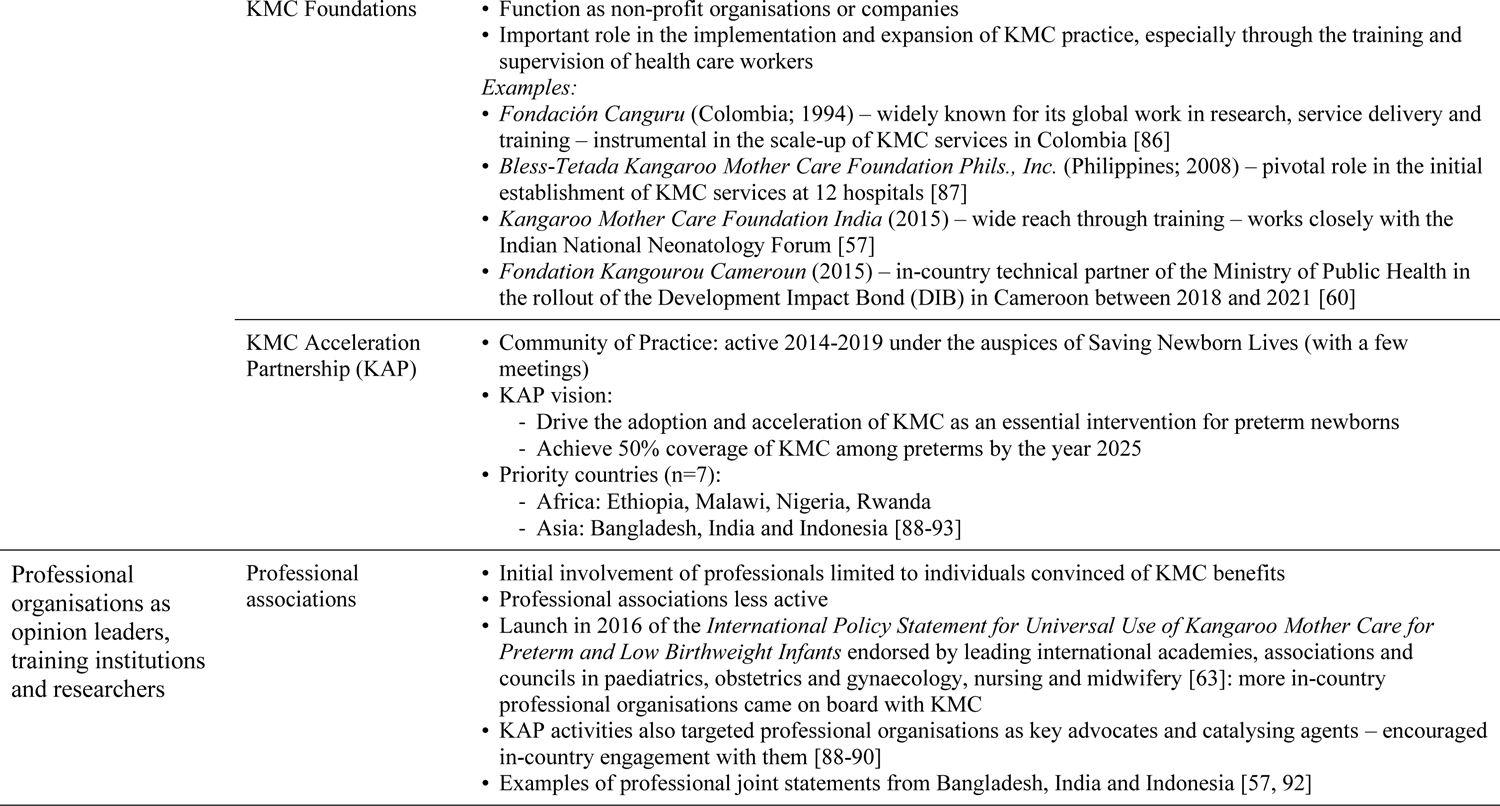
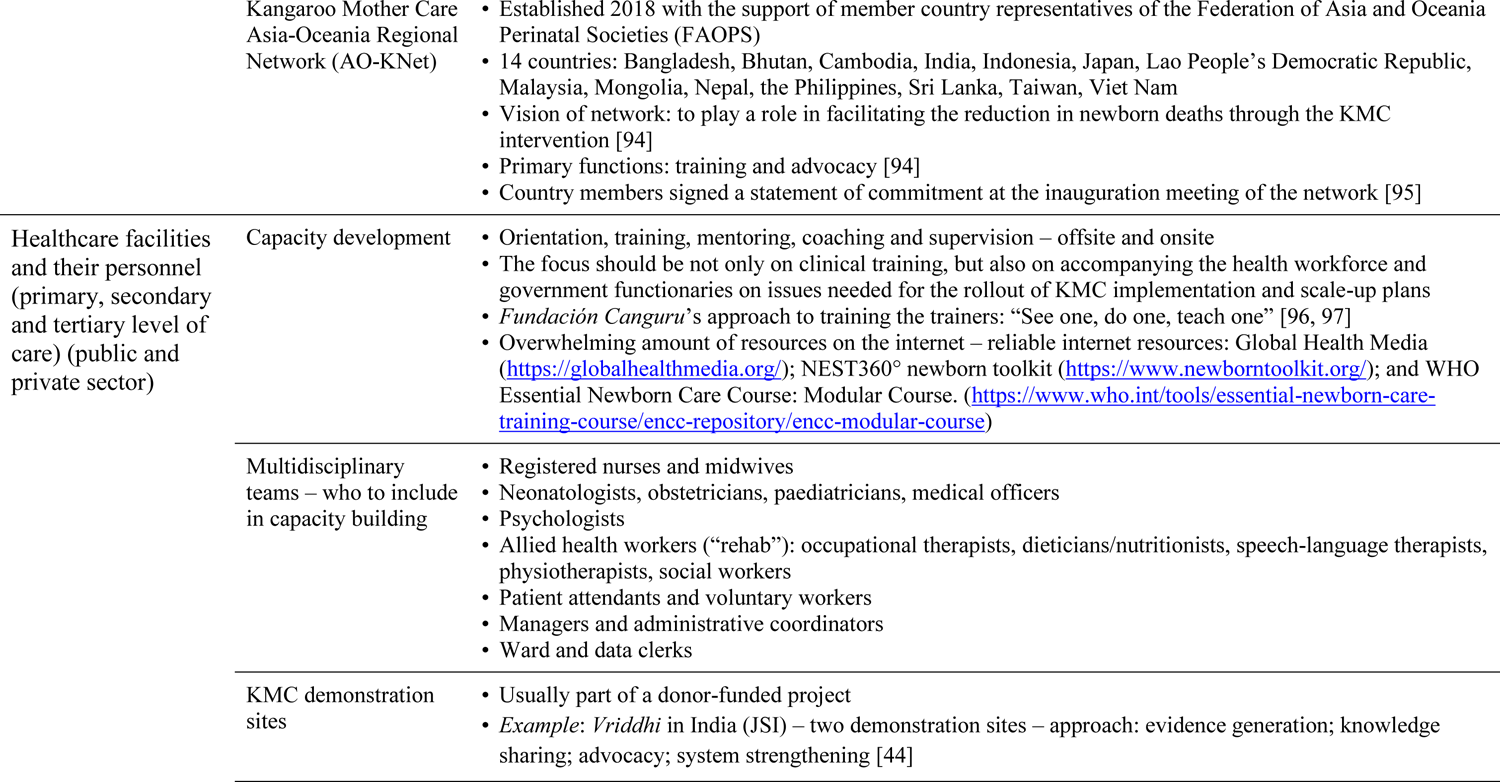
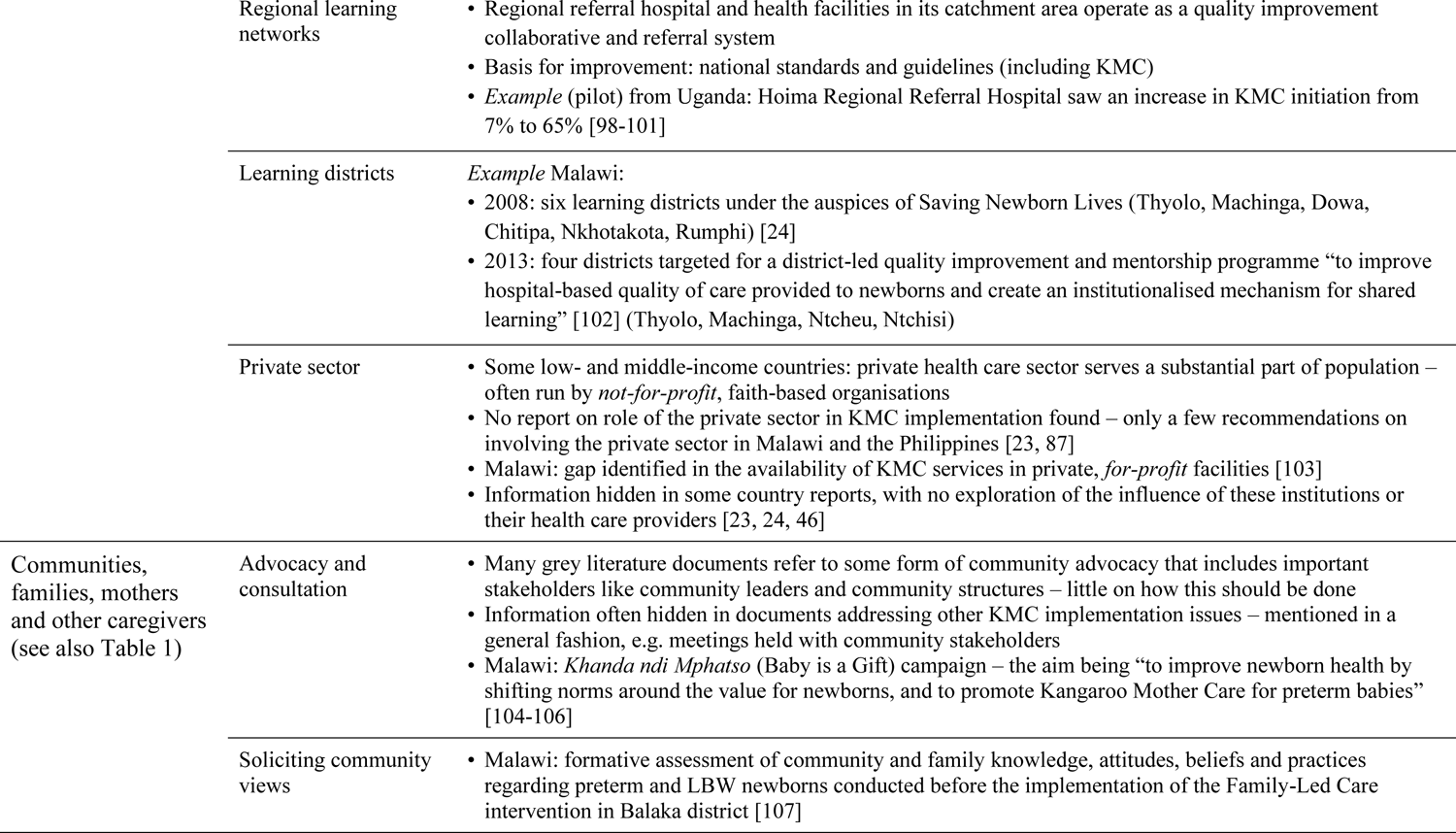
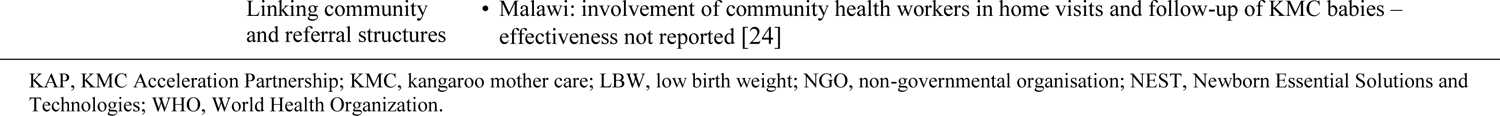
Description of implementing partners

#### Process: initial pathways to KMC implementation and scale-up

Originally KMC was introduced at national, subnational or health-facility level in different fashions, with KMC services mostly introduced – after exposure to KMC [16, 53] – by committed and personally motivated professionals in individual health facilities. At the Istanbul KMC Acceleration Convening in 2013, De Graft-Johnson referred to a number of programmatic approaches to the introduction and expansion of KMC services [9], which are summarised in Table 3 with further explanations and examples.

**Table 3.** Programmatic approaches to the introduction and expansion of KMC services

| **Approach [9]** | **Explanation and examples** |
| --- | --- |
| One institution to another (“institutional teams learning abroad” [108]) | The Colombian model of training teams from other countries – the principle of “See one, do one, teach one” [96, 97] |
| Vertical Phase-in approach with support from international NGOs/donors in partnership with MOH | Much of the early work in Africa and India |
| Local KMC Foundation led expansion with accreditation | The Philippines KMC Foundation (see Table 2) |
| Integrated approach – KMC training a component of WHO ENC package, country IMNCI and BEmONC packages, with or without “pedestal” support | “Pedestal support” refers to additional support specifically for KMC [109, 110] – this was the approach used in a Ghana KMC scale-up initiative |
| Regional Alliance approach | The Latin American and Caribbean (LAC) Neonatal Alliance promoted KMC [45, 111] |
| Provincial Health Authority approach | Technical assistance to subnational authorities |

BEmONC, Basic Emergency Obstetric and Newborn Care; ENC, Essential Newborn Care; IMNCI, Integrated Management of Newborn and Childhood Illness; KMC, kangaroo mother care; MOH, Ministry of Health; NGO, non-governmental organisation; WHO, World Health Organization.

We constructed a crude categorisation in an attempt to understand the history of KMC implementation and scale-up over more than three decades (Figure 3). There were two main pathways of implementation and scale-up, both starting with some form of KMC practice at individual hospitals. One pathway emerged from within the public health system with donor support channelled through government structures. Brazil [20, 21] and South Africa [25, 26] are examples of this pathway. In 2000, Brazil included KMC in its *Sistema Único de Saúde* package of medical assistance paid for by the government [20] whereas since 1994 South Africa has provided free public health care services to mothers and children under five years of age [56]. The second pathway included an intermediate phase where an external driver provided technical support for KMC implementation, often in a small group of health facilities or in one or more districts in a country. This corresponds to the Vertical Phase-in approach mentioned in Table 3.

**Figure 3.**
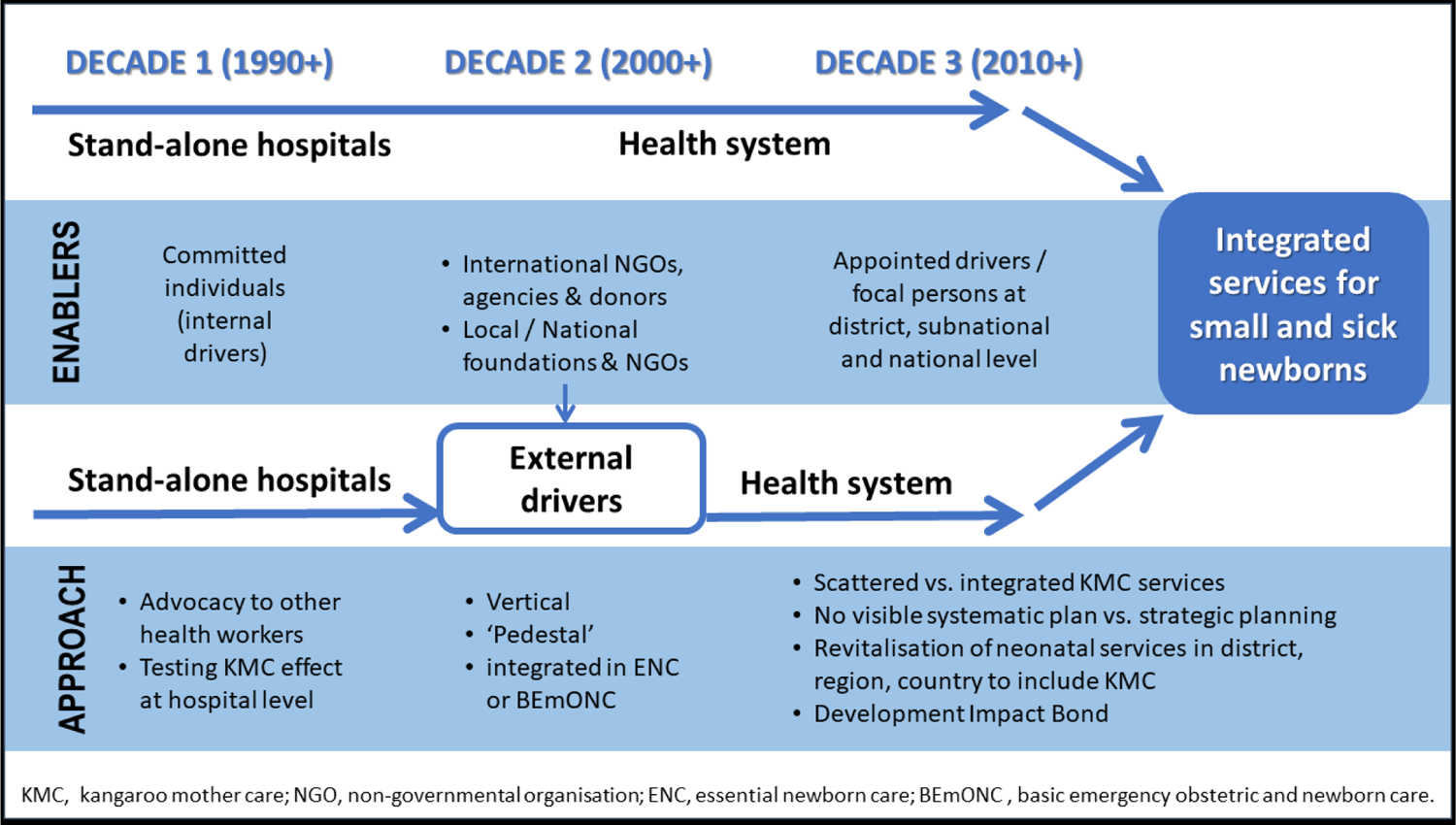
Broad categorisation of trajectories of KMC implementation.

External drivers serving as catalysts for KMC implementation included international non-governmental organisations, agencies and donors. Save the Children through its Saving Newborn Lives programme was one of the agencies that were active in many countries. Malawi is the country that appears to have received the most external support over more than two decades and KMC space and units have expanded rapidly there. Despite this expansion, population-based coverage is still low.

In Colombia and the Philippines a national KMC foundation acted as an external driver in the expansion of in-country KMC services. Their roles are described in detail in Table 2. The Indian KMC Foundation supports the government of the country in implementing KMC and is active in organising annual conferences to showcase research and experience in different clinical and implementation aspects of KMC [57]. A world first for KMC was the scale-up of KMC to 10 hospitals across Cameroon through a Development Impact Bond (DIB) [58]. The *Fondation Kangourou Cameroun* led the in-country implementation with the support of the Colombian Foundation. The model is data-driven, providing for rigorous performance management, but also allowing for flexibility in the adaptation of KMC service delivery to the local context of individual health facilities [59, 60].

Some countries adopted a more holistic and integrated programmatic approach to KMC implementation. For example, in Rwanda newborn care improvements are coordinated by the Ministry of Health in Rwanda and KMC implementation was embedded in the revitalisation of neonatal units in district hospitals. Collaboration between partners enabled the procurement of equipment, the development of national KMC guidelines and newborn protocols, coordinated training and formative supervision [61].

## DISCUSSION

Exploring the synthesis of grey literature on the programmatic implementation of KMC involved a journey through uncharted territory. Despite the large volume of documents screened, sifted and analysed, few reports on population-based KMC and the national or subnational availability of KMC services were found. Health system factors involved in the programmatic implementation of KMC were identified and the lessons learnt were organised along the lines of the health system building blocks. Contextual factors were divided into people as catalysts for change and pathways initially followed in the implementation of KMC.

In the grey literature, information on efforts to programmatically scale up KMC services at national and subnational level was available for 18 low- and middle-income countries. In most of these countries there is a strong presence of development agencies with a focused maternal and newborn health agenda. With regard to uncoordinated and scattered implementation, Aliganyira et al observed “a highly inequitable distribution of KMC services” in Uganda (p6) [62]. This points to the urgent need to collect and analyse data on KMC-related indicators at subnational level to ensure equity of intervention impact in the most vulnerable populations.

Possible explanations for the dearth of data on national KMC services from higher-income countries include health system differences and the emphasis on the autonomy of clients. It appears as if the implementation of some form of KMC practice is driven by members of professional associations in the individual health facilities that incorporate KMC into their protocols for small and sick newborns without seeing a need to publicise a KMC service or collect and analyse KMC-specific data for individual patients. The launching of the *International Policy Statement for Universal Use of Kangaroo Mother Care for Preterm and Low Birthweight Infants* [63] in 2016 can be seen as an attempt to leverage the influence of professional associations in the acceleration of KMC implementation. A systematic review and realist synthesis of KMC scale-up in the United Kingdom identified lack of training, knowledge and confidence, and absence of clear guidelines as the main barriers. The authors propose a focus on the potential cost-effectiveness of KMC in reducing the need for incubator use [64].

Overall, the “what” and “how” of KMC implementation and scale-up could be equated to the capacity development needed to implement a national or subnational KMC programme. According to the United Nations Development Programme, capacity development is a process of change that could include various types of education, training and workshopping activities. Capacity development also refers to the strengthening of the health system to improve performance and ensure sustainability [65].

The health system factors influencing the programmatic implementation of KMC are also reported on in the published literature in terms of health system building blocks [11, 45, 66–68]. One issue emerging from the review is the challenge of establishing a continuum of care through linking the health facility with the community and changing negative community and family perceptions around preterm birth that may influence the uptake of and adherence to KMC, especially after discharge from the health facility. These issues are also reflected in a number of systematic and scoping reviews on barriers to and enablers of the adoption of KMC [5, 7, 69, 70]. Achieving intensive involvement of community health workers who pay frequent, regular home visits still seems to be a hurdle in many communities [71], although studies by Mazumder et al [72] and Mony et al [11] have respectively shown that better health outcomes and high KMC coverage are achieved where there are frequent visits by community health workers.

The context of programmatic KMC implementation is navigated by the people driving the process. This is linked to the health system building block of leadership and governance, within the health system and beyond. Of particular importance is the way successful people working as individuals and in teams can leverage change and create a potentially sustainable programme. Many of the documents in the grey literature were produced by implementing partners described in Table 2 as donors, development agencies and international non-governmental organisations. The challenges of fragmentation and “end-of-project syndrome” (i.e. end of funding) associated with this category of implementing partners are captured by Spicer et al in a publication titled: “The development sector is a graveyard of pilot projects!” [73]. They also found that innovations must be perceived as effective, require modest resource inputs, be acceptable to and incentivise frontline health workers, be acceptable to communities, and be adaptable across diverse geographical contexts [73]. These conditions could also be applied to the programmatic implementation of KMC in the design of implementation and scale-up plans and in advocacy messages.

Capacity development of the health workforce at all levels is indispensable in any KMC implementation. These levels include the public health leaders, administrators and programme managers at national and subnational level and personnel at health facility and community level. Initially, when KMC was still considered a novel intervention, stand-alone training in KMC was often the order of the day [74]. A challenge in the current environment where KMC is included in the training packages for the care of small and sick newborns is to ensure that different approaches to the organisation of KMC services and different types of knowledge and skills needed for different levels of facilities are accounted for responsibly.

The categorisation of three decades of KMC implementation and scale-up in Figure 3 is similar to the three waves of KMC implementation as described in a case study on three Asian countries [74]. KMC implementation and expansion are currently far more deeply embedded in the upgrading and scale-up of neonatal services. In a study conducted on the scale-up of maternal and newborn health innovations in Ethiopia, India and Nigeria, Spicer and colleagues concluded “that scale-up has no magic bullet solution – implementers must embrace multiple activities, and require substantial support from donors and governments in doing so” (p30) [19]. In Table 4 we use a stages-of-change framework initially developed for monitoring progress with KMC implementation [75, 76] to reconstruct a number of possible actions for scaling up a national KMC programme, with a special focus on the health system and health facilities. For each action, governments should find contextually appropriate answers and solutions [11]. Some actions pertain to KMC specifically; others should be aligned with the newborn health agenda and programme.

**Table 4.** What is needed to scale up a national KMC programme?

| STAGE OF CHANGE | ACTION | DESCRIPTION OF ACTIVITIES |
| --- | --- | --- |
| 1. Create awareness | Advocacy | • Identify advocacy needs [112] and adopt effective approaches to advocacy for KMC [19]<br>• Generate and present strong evidence to support government decision making to adopt and finance a KMC programme [19, 36, 73]<br>• Promote community uptake of KMC through involvement of media, community leaders, mobilisation teams and role models [19, 27, 36]<br>• Promote male involvement in the care of the small newborn [36, 113, 114]<br>• Use opinion leaders at various levels of the health system, in professional associations and in the community to promote KMC [36, 110, 115] |
| 2. Commit to implementation at scale | Leadership | • Ensure the Ministry of Health is the leader or primary stakeholder of the implementation process [60, 111], responsible for developing KMC policies and a KMC programme [10, 19, 27]<br>• Endorse KMC in important national neonatal policy and strategic documents [10] |
|  | Programming | • Contextualise the implementation strategies [12] and embed scale-up in a shared programme design that: is data driven [8, 78]; has clear and feasible objectives [78, 111] and appropriate communication and networking strategies [116, 117]; allocates time and resources [19, 73]; and has a costing plan for the expansion of KMC services [36] that include measures for ensuring the provision of equitable KMC services in the least-resourced districts or facilities with the highest numbers of deliveries [36, 62]<br>• Align programming with other MNCH initiatives like the BFHI [61, 118] and EmONC activities [112]<br>• Strengthen harmonisation and align activities among external programmes [19] (including any in-country performance-based frameworks) [115] to avoid fragmentation and promote government ownership [78, 119] – also conduct stakeholder consultations [108]<br>• Get the private sector health system and facilities on board [11, 23, 87] |

**Table 4.** What is needed to scale up a national KMC programme?
| STAGE OF CHANGE | ACTION | DESCRIPTION OF ACTIVITIES |
| --- | --- | --- |
|  | Manage funding | • Endeavour to reduce donor dependency [47] and be creative in leveraging various funding sources [29]<br>• Secure longer-term funding from donors and development partners to minimise uncertainty [73]<br>• Work towards limiting out-of-pocket expenditure for families as far as possible [115, 120] and including KMC in national health insurance care packages [29, 121]<br>• Be creative in leveraging funding |
| 3. Prepare for implementation at scale | Planning and leadership | • Establish multiple stakeholder task teams or steering committees consisting of committed experts (including national champions for KMC and representatives of professional associations) at national, subnational and facility level as needed [10, 36, 82, 108, 111, 115]<br>• Create sustainable business plans that include human resource, financial and material needs [112, 122] and reliable performance measurements [78]<br>• Align communication channels, linkages and responsibilities among policymakers, different health facilities with different owners and different line managers (e.g. public [central vs. district], faith-based, private for profit) [23] and health-care users [112] |
|  | Assessment and analysis | • Conduct a KMC service readiness assessment at subnational entities and health facilities [66, 112, 115, 123, 124] or some form of bottleneck [45, 118, 125] or situation analysis [126-129], including a survey on the quality of KMC presence in pre-service health curricula and training programmes [66, 129], to inform the details of implementation and scale-up plans |

**Table 4.** What is needed to scale up a national KMC programme?
| STAGE OF CHANGE | ACTION | DESCRIPTION OF ACTIVITIES |
| --- | --- | --- |
|  | Strengthening health systems | • Harness the resources, infrastructure and capacity of existing MNCH programmes [110] and embed and ensure full integration of KMC into these programmes [8, 47, 77, 115, 130]<br>• Integrate KMC indicators into the district and national HIMS (e.g. DHIS2) [8, 110, 131], with KMC-specific indicators on coverage and quality [11, 132-134] and KMC service outcomes as a district performance indicator [36]<br>• Strengthen health systems [19] and core neonatal infrastructure [115], and support individual health facilities with changes in infrastructure and equipment to enable the provision of quality KMC services [10, 11, 29, 33, 36, 45, 47, 85, 87, 108, 110, 120, 131, 135-143]<br>• Identify or develop one or more centres of excellence [10, 45, 60, 74, 109, 110, 115, 131, 135, 144-146]<br>• Strengthen referral systems where needed [143] |
|  | Building capacity | • Build implementer capacity to improve service delivery [60, 115, 125] and catalyse scale-up [19, 147]<br>• Provide high-quality training to multiple cadres of facility staff and community health workers through the use of facilitators with a background and practical experience in KMC [23, 79, 108, 115, 129]<br>• Develop a plan for outreach and supportive supervision and allocate responsibilities to specific role players [36] |
|  | Sensitising and learning | • Sensitise health care service providers and subnational managers about the introduction of KMC [125, 147] to minimise provider hesitancy [12, 125]<br>• Develop or adapt suitable training material [10]<br>• Develop or adapt BCC materials and distribute them to health facilities and communities [12, 143, 147, 148] |

**Table 4.** What is needed to scale up a national KMC programme?
| STAGE OF CHANGE | ACTION | DESCRIPTION OF ACTIVITIES |
| --- | --- | --- |
|  | Service delivery | • Use an action sequence to plan for the delivery of KMC services: identify small babies; initiate KMC per protocol; continue KMC to discharge; and follow up KMC babies to KMC “graduation” (discharge from KMC) [66]<br>• Design protocols and SOPs for KMC [36, 115, 128] as part of the guidelines developed for the care of small and sick newborns that can be adapted at health facility level – promote ownership of centrally-designed documents at subnational and facility level [36] |
|  | Monitoring and evaluation | • Investigate how KMC indicators could be included in existing records and determine whether any new standardised records or registers should be developed at the national and/or subnational level [120]<br>• Develop HMIS tools, e.g. monthly report forms [41] and train designated staff at all levels of the health system on data capturing, analysis, sharing and use of results [36, 134] |
| 4. Implement KMC in facilities and across subnational entities | Service delivery | • Provide clear guidance to the health workforce and other stakeholders on expected roles and responsibilities with regard to KMC implementation at health-facility and community level and the scale-up of KMC in a particular administrative region [36, 61, 110, 112]<br>• Adapt service delivery to local contexts [60] and build in flexibility in implementation [11, 78]<br>• Strengthen staff [19], minimise staff rotations [12, 36, 129] and mitigate the effect of clinical staff turnover [60, 115] |
|  | Documentation | • Adapt national and subnational KMC protocols and SOPs at health facility level [110, 115] and implement all the components of KMC: position, nutrition, discharge and follow-up, as well as support [1, 3, 4, 120]<br>• Implement the national or subnational records and registers and promote accuracy in entering information and data |

**Table 4.** What is needed to scale up a national KMC programme?
| STAGE OF CHANGE | ACTION | DESCRIPTION OF ACTIVITIES |
| --- | --- | --- |
|  | Learning | • Do systematic outreach to lower level health facilities and provide structured supportive supervision within the health system that includes onsite coaching and mentoring [29, 36, 60, 144, 149] and clinical shadowing [115]<br>• Promote strong communities of practice [115] and learning networks between healthcare workers, facilities and administrative structures [98] |
|  | Focusing on healthcare users | • Review and change hospital procedures to provide family-centred services (e.g. 24-hour visiting policies and accommodation for parents/families) [67, 117, 146] and to allow the mother to have a companion of choice to assist with KMC [36, 110]<br>• Ensure that all mothers, families and other caregivers in KMC receive sufficient counselling on KMC and the danger signs [60]<br>• Establish strong health facility–community linkages [11], which include campaigns [143, 148] and community education activities [29] |
| 5 Integrate KMC into routine practice and health systems activities | Service delivery | • Maintain an effective referral and follow-up system at subnational level [12, 66, 115]<br>• Integrate KMC into quality-of-care initiatives at facility and subnational level [12, 150] |
|  | Documentation | • Maintain high-quality record keeping [110] |
|  | Learning | • Provide regular in-service orientations for novices and new staff and refresher activities for all staff at health-facility and community level [12, 36, 112, 115], also with regard to improving data quality and use of data [134] |
|  | Monitoring and evaluation | • Monitor and support the performance of staff and health facilities [11, 143] with built-in accountability mechanisms [129]<br>• Maintain regular, standardised reporting to the HMIS [110, 148] and use the data to identify and support struggling districts and individual facilities [36] |

**Table 4.** What is needed to scale up a national KMC programme?
| STAGE OF CHANGE | ACTION | DESCRIPTION OF ACTIVITIES |
| --- | --- | --- |
| 6 Sustain KMC practice at scale | Learning | • Continue support for induction of new staff, refreshers and mentorship and coaching activities [112]<br>• Sustain learning networks among healthcare workers, facilities and administrative structures |
|  | Monitoring and evaluation | • Sustain high-quality recordkeeping, regular reporting to the health information system and use of data [29, 110] |
|  | Assessment and research | • Measure implementation fidelity and sustainability against the original business plan [12, 122] and analyse the cost of embedding KMC in neonatal units [115]<br>• Operationalise success [78], document the implementation process as a learning opportunity and combine it with implementation research to understand what else would be needed to reach sustainable practice [11, 68, 72, 120] |

BCC, behaviour change communication; BFHI, Baby-Friendly Hospital Initiative; DHIS2, District Health Information Software 2; HMIS, health management information system; KMC, kangaroo mother care; EmONC, emergency obstetric and newborn care; MNCH, maternal, newborn and child health; SOP, standard operating procedure.

### Study limitations

This review illustrated how hard it is to find relevant information on an intervention that is embedded in a broader package of small and sick newborn care. Important documents may also have been missed because not all languages were accommodated. Governments and development partners in some countries seem to be more forthcoming than others regarding the documents they place on the internet and their activities naturally received more attention in the review. The reviewer’s involvement in the assessment of KMC services of many countries has influenced the analysis, but has also enabled the inclusion of documents that might otherwise not have been found. Lastly, the review did not include an in-depth analysis of how certain cross-cutting themes featured in the programmatic implementation of KMC because of the large volume of documents and uncertainty about how these themes could be identified with common search terms. Broader themes derived from the ENAP’s guiding principles are examples of such issues for future attention: human rights; integration; equity; accountability; and innovation [77]. Some of the information summarised in this review has also found its way into peer-reviewed publications, which were not covered by this study.

## CONCLUSION

The synthesis of the grey literature contributes to the understanding of how programmatic implementation of KMC takes place and which aspects could be considered in the design and scale-up of such programmes. Two of the main takeaways from this review are: avoid “empty scale-up” with an “appearance of success” [78], and “balance rapid expansion of services with the need to improve quality of care” [36] for small and sick newborns. Programmatic implementation of KMC requires action in multiple health system building blocks with a focus on monitoring and evaluation of availability and quality of services along with coverage.

## Data Availability

Materials are available upon reasonable request from the corresponding author.

## Acknowledgments

We are grateful to individuals, governments and organisations who shared documents not available on the internet, to Karen Edmond for her valuable inputs into the conceptualisation of the review and to Nils Bergman for reviewing the original work plan.

## REFERENCES

1. Conde-Agudelo A, Diaz-Rossello JL. Kangaroo mother care to reduce morbidity and mortality in low birthweight infants. Cochrane Database Syst Rev. 2016(8):Cd002771.

2. World Health Organization. WHO recommendations on interventions to improve preterm birth outcomes. Geneva: World Health Organization; 2015.

3. World Health Organization. Kangaroo mother care: a practical guide. Geneva: World Health Organization; 2003.

4. Nyqvist K, Anderson C, Bergman N, Cattaneo A, Charpak N, Davanzo R, et al. State of the art and recommendations: Kangaroo mother care: application in a high-tech environment. Acta Paediatr. 2010;99:812–9.

5. Smith ER, Bergelson I, Constantian S, Valsangkar B, Chan GJ. Barriers and enablers of health system adoption of kangaroo mother care: a systematic review of caregiver perspectives. BMC Pediatr. 2017;17(1):35.

6. Chan G, Bergelson I, Smith ER, Skotnes T, Wall S. Barriers and enablers of kangaroo mother care implementation from a health systems perspective: a systematic review. Health Policy Plan. 2017;32(10):1466–75.

7. Seidman G, Unnikrishnan S, Kenny E, Myslinski S, Cairns-Smith S, Mulligan B, et al. Barriers and enablers of kangaroo mother care practice: a systematic review. PloS One. 2015;10(5):e0125643.

8. World Health Organization, United Nations Children’s Fund. Survive and thrive: transforming care for every small and sick newborn. Geneva: World Health Organization; 2019.

9. de Graft-Johnson J. Introduction and Expansion of Kangaroo Mother Care Services: Programmatic Approaches (unpublished presentation). KMC Acceleration Convening, Istanbul, Turkey, 21–22 October 2013.

10. Torres LM, Mazia G, Guenther T, Valsangkar B, Wall S. Monitoring the implementation and scale-up of a life-saving intervention for preterm and small babies: Facility-based Kangaroo Mother Care. J Glob Health. 2021;11:14001.

11. Mony PK, Tadele H, Gobezayehu AG, Chan GJ, Kumar A, Mazumder S, et al. Scaling up Kangaroo Mother Care in Ethiopia and India: a multi-site implementation research study. BMJ Glob Health. 2021;6(9):e005905.

12. Mehjabeen S, Matin M, Gupta RD, Sutradhar I, Mazumder Y, Kim M, et al. Fidelity of kangaroo mother care services in the public health facilities in Bangladesh: a cross-sectional mixed-method study. Implement Sci Commun. 2021;2(1):115.

13. Kumar H, Bhat A, Alwadhi V, Maria A, Khanna R, Neogi SB, et al. An assessment of implementation of family participatory care in special newborn care units in three states of India. Indian Pediatr. 2021;58(4):349–53.

14. World Health Organization. Monitoring the building blocks of health systems: a handbook of indicators and their measurements strategies. Geneva: World Health Organization; 2010.

15. Glasgow RE, Vogt TM, Boles SM. Evaluating the public health impact of health promotion interventions: the RE-AIM framework. Am J Public Health. 1999;89(9):1322–7.

16. Maternal and Child Health Integrated Program (MCHIP), USAID. Kangaroo mother care implementation guide. Washington, D.C.: MCHIP; 2012. Available: https://www.mchip.net/technical-resource/kangaroo-mother-care-implementation-guide/. Accessed: 7 October 2021.

17. CORE Group. “SCALE” and “SCALING-UP”: A CORE Group Background Paper on “Scaling-Up” Maternal, Newborn and Child Health Services. 11 July 2005. Available: https://www.mchip.net/sites/default/files/scaling_up_background_paper_7-13.pdf. Accessed: 5 October 2021.

18. QSR International Pty Ltd. NVivo (Version 9). 2010. Available: https://www.qsrinternational.com/nvivo-qualitative-data-analysis-software/home. Accessed: 20 October 2021.

19. Spicer N, Bhattacharya D, Dimka R, Fanta F, Mangham-Jefferies L, Schellenberg J, et al. ’Scaling-up is a craft not a science’: Catalysing scale-up of health innovations in Ethiopia, India and Nigeria. Soc Sci Med (1982). 2014;121:30–8.

20. Gómez HM. Kangaroo Mother Care. English version. Brazilian Development Bank, Social Operations Department; ca 2000. Available: https://www.bndes.gov.br/SiteBNDES/export/sites/default/bndes_en/Galerias/Download/studies/KangarooMother.pdf. Accessed 12 November 2021.

21. Lamy ZC, Geaquinto L. Kangaroo care coverage: from pilot projects to scale: Brazil (presentation). XI Workshop and Congress of the International Network on Kangaroo Mother Care, Trieste, Italy, 14–17 November 2016. Available: http://fundacioncanguro.co/wp-content/uploads/2018/01/16_lamy_machado_KMC-Brazil.pdf. Accessed: 23 July 2021.

22. Brasil Ministério da Saúde. Atenção Humanizada ao Recém-Nascido: Método Canguru. Manual Técnico [Attention to Humanized Newborn Care: Kangaroo Method. Technical Manual]. (3rd ed). Brasilia: Ministério da Saúde, Departamento de Ações Programáticas Estratégicas; 2017. Available: https://bvsms.saude.gov.br/bvs/publicacoes/atencao_humanizada_metodo_canguru_manual_3ed.pdf. Accessed: 4 August 2021.

23. Bergh A-M, Van Rooyen E, Lawn J, Zimba E, Ligowe R, Chiundu G. Retrospective evaluation of kangaroo mother care practices in Malawian hospitals, July-August 2007. Report. Lilongwe: Malawi Ministry of Health; 2007. Available: http://www.healthynewbornnetwork.org/hnn-content/uploads/SNL-2007.-Malawi-KMC-Assessment-Report.pdf. Accessed: 20 July 2021.

24. Bergh A-M, Banda L, Lipato T, Ngwira G, Luhanga R, Ligowe R. Evaluation of kangaroo mother care services in Malawi, February 2012. Research report. Ministry of Health, Save the Children, USAID/MCHIP; 2012. Available: http://www.healthynewbornnetwork.org/hnn-content/uploads/8-Malawi-2012-KMC-Assessment-report.pdf. Accessed: 20 July 2021.

25. Perinatal Priorities. Website of the Priorities in Perinatal Care Association of South Africa (with proceedings and abstracts). Available: https://www.perinatalpriorities.co.za/. Accessed: 20 August 2021.

26. MRC Unit for Maternal and Infant Health Care Strategies, PPIP Users, National Department of Health. Saving Babies 2002: Third Perinatal Care Survey of South Africa Pretoria: MRC Unit for Maternal and Infant Health Care Strategies; 2002. Available: https://www.up.ac.za/media/shared/717/PPIP/Saving%20Babies%20Reports/report-3-saving-babies-2002.zp194943.pdf(Saving. Accessed: 24 August 2021.

27. Hailegebriel TD, Bergh AM, Zaka N, Roh JM, Gohar F, Rizwan S, et al. Improving the implementation of kangaroo mother care. Bull World Health Organ. 2021;99(1):69–71.

28. Charpak N, Lince C. KMC mapping in Colombia: An example of KMC diffusion at country scale (unpublished presentation). XIII Workshop and Congress of the International Network on Kangaroo Mother Care, Madrid, Spain, 21–24 November 2022.

29. Calibo AP, De Leon Mendosa S, Silvestre MA, Murray JCS, Li Z, Mannava P, et al. Scaling up kangaroo mother care in the Philippines using policy, regulatory and systems reform to drive changes in birth practices. BMJ Glob Health. 2021;6:e006492.

30. Ehtesham Kabir A, Afroze S, Amin Z, Biswas A, Lipi SA, Khan M, et al. Implementation research on kangaroo mother care, Bangladesh. Bull World Health Organ. 2022;100(1):10–9.

31. Chavula K. Improving Availability, Quality, and Use of Routine date for Newborns: Malawi’s experience with kangaroo mother care. Save the Children’s presentation on the Kangaroo Mother Care initiative in Malawi. 29th International Pediatric Association Congress, Panama City, Panama, 17-21 March 2019. Available: https://resourcecentre.savethechildren.net/library/improving-availability-quality-and-use-routine-date-newborns-malawis-experience-kangaroo. Accessed: 27 October 2021.

32. Viet Nam Ministry of Health, MCH Dept., KMC team Viet Nam. Kangaroo mother care in Viet Nam: an overview for implementation and scaling up (abstract). XI Workshop and Congress of the International Network on Kangaroo Mother Care, Trieste, Italy, 14-17 November 2016. Available: https://fundacioncanguro.co/wp-content/uploads/2018/01/vietnam.docx. Accessed: 12 June 2022.

33. Bergh A-M, Van Rooyen E. Survey of kangaroo mother care in North West Province, 2015 (unpublished presentation).; Maternal, Child and Women’s Health Meeting, Mahikeng, South Africa. 23 February 2016.

34. Vakilian R, Heidarzadeh M, Akrami F, Ravari M, Habibelahi A, Khazaei S, et al. Development of a national service package for the implementation of kangaroo mother care in Iran (abstract). lX lnternational Conference on Kangaroo Mother Care, Ahmedabad, India, 22–25 November 2012. Available: https://fundacioncanguro.co/documentacion-canguro/. Accessed: 23 June 2021.

35. Morales Betancourt P, López Maestro M, De la Cruz Bértolo J, Vázquez Roman S, Alonso Díaz C, Pallás Alonso C. What we talk about in NICUs when we talk about kangaroo care: a national survey in Spain (abstract). XII International Conference on Kangaroo Mother Care, Bogotá, Colombia, 14–17 November 2018. Available. http://fundacioncanguro.co/wp-content/uploads/2018/11/What-we-talk-about-in-NICUs-When-we-talk-about-Kangaroo-Care.-A-National-Survey-in-Spain.pdf. Accessed: 20 July 2021.

36. Save the Children, USAID/MCHIP, MRC University of Pretoria. Tracking implementation progress of Kangaroo Mother Care. Overview of results of a multi-country evaluation (policy brief). 2014. Available: https://www.healthynewbornnetwork.org/resource/tracking-implementation-progress-for-kangaroo-mother-care-overview-of-results-from-a-multi-country-evaluation/. Accessed: 26 October 2021.

37. World Health Organization, United Nations Children’s Fund. Every Newborn progress report 2019. Geneva: World Health Organization; 2020.

38. Charpak N, Villegas J. Colombia. Working group contribution. XI Workshop and Congress of the International Network on Kangaroo Mother Care, Trieste, Italy, 14–17 November 2016. Available: https://fundacioncanguro.co/wp-content/uploads/2018/01/Colombia.pdf. Accessed: 30 June 2021.

39. Charpak N, Godoy N. Estrategia Programas Madre Canguro en Colombia: Antecedentes y Perspectivas [Kangaroo Mother Care Program Strategy in Colombia: Background and Perspectives] (unpublished presentation). IV National Workshop on the Kangaroo Mother Care Method, Bogotá, Colombia, 24 January 2020.

40. Save the Children Malawi. Improving availability and quality of routine data for newborns: Malawi’s experience with kangaroo mother care (policy brief). Lilongwe, Malawi: Save the Children; 2018. Available: https://www.healthynewbornnetwork.org/resource/improving-routine-data-for-newborns-malawi-experience/. Accessed: 28 May 2020.

41. Torres LM, Mazia G, Guenther T, Valsangkar B, Wall S. Monitoring the implementation and scale-up of a life-saving intervention for preterm and small babies: Facility-based Kangaroo Mother Care (Online Supplemementary Document). J Glob Health. 2021;11:14001. http://www.jogh.org/documents/2021/jogh-11--s001.pdf.

42. Mugeni C, Kayinamura Mwali A. Ensuring Quality of Care for Small Babies in Rwanda (presentation). XI Workshop and Congress of the International Network on Kangaroo Mother Care, Trieste, Italy, 14–17 November 2016. Available: https://fundacioncanguro.co/wp-content/uploads/2018/01/19_mugeni_rwanda.pdf. Accessed: 23 July 2021.

43. Vani SN, Banker D. Enablers and Challenges of Kangaroo Mother Care practices at different levels of newborn care services in India. Document prepared for a working group. XI Workshop and Congress of the International Network on Kangaroo Mother Care, Trieste, Italy, 14–17 November 2016. Available: https://fundacioncanguro.co/wp-content/uploads/2018/01/india.docx. Accessed: 23 July 2021.

44. Vriddhi, USAID. JSI’s contribution under Vriddhi for implementation of Kangaroo Mother Care in District Hospitals. ca. 2017. Available: https://www.jsi.com/JSIInternet/Inc/Common/_download_pub.cfm?id=19256&lid=3. Accessed: 24 July 2021.

45. Vesel L, Bergh A-M, Kerber KJ, Valsangkar B, Mazia G, Moxon SG, et al. Kangaroo mother care: a multi-country analysis of health system bottlenecks and potential solutions. BMC Pregnancy Childbirth 2015;15(Suppl 2):S5.

46. Bergh A-M, Davy K, Otai CD, Nalongo AK, Sengendo NH, Aliganyira P. Evaluation of kangaroo mother care services in Uganda. Save the Children, USAID/MCHIP; 2012. Available: https://www.healthynewbornnetwork.org/hnn-content/uploads/13-Uganda-2012-KMC-review-report.pdf. Accessed 10 November 2021.

47. Bergh A-M, Bello Mohammed S, Vaz L, Oyinbo M, Omale L, Madubuko B, et al. Public hospital-based care of small newborns in Nigeria. Research report. Abuja: Save the Children; 2017. Available: https://www.healthynewbornnetwork.org/hnn-content/uploads/NigeriaCareForSmallNewborns.pdf. Accessed: 28 September 2021.

48. Medvedev MM. Informing the design of a trial of kangaroo mother care initiated before stabilisation amongst small and sick newborns in a sub-Saharan African context using mixed methods: PhD thesis, London School of Hygiens and Tropical Medicine; 2020. Available: https://researchonline.lshtm.ac.uk/id/eprint/4658155/. Accessed: 26 June 2021.

49. Misra A, Cunningham S. Kangaroo Mother Care—High on Evidence, Low on Reach (brief). JSI. 2017. Available: https://www.jsi.com/kangaroo-mother-care-high-on-evidence-low-on-reach/. Accessed: 24 July 2021.

50. Maternal and Child Survival Program (MCSP)/USAID. The feasibility and acceptability of Kangaroo Mother Care in the neonatal unit of Women and Children’s Hospital, Taunggyi, Myanmar Study Report. 2018. Available: https://resourcecentre.savethechildren.net/library/feasibility-and-acceptability-kangaroo-mother-care-neonatal-unit-women-and-childrens. Accessed: 27 July 2021.

51. de Graft-Johnson J, Wall S, Daly P, Mwebesa W, Kerber K. Lessons learned from the introduction and expansion of Kangaroo Mother Care services in selected developing countries (poster). Save the Children. 2007. Available: https://www.healthynewbornnetwork.org/resource/lessons-learned-from-the-introduction-and-expansion-of-kangaroo-mother-care-services-in-selected-developing-countries/. Accessed: 25 July 2021.

52. Unnikrishnan S. Introduction to breakouts – Approaches for KMC acceleration (slides 130-136) (unpubilshed). KMC Acceleration Convening, Istanbul, Turkey, 21–22 October 2013.

53. Bergh A-M, Hoque D, Udani R, Rao S, Puri A, Pratomo H, et al. The implementation and scale up of facility-based kangaroo mother care in five Asian countries, September 2014. Unpublished research report. USAID/MCHIP and Save the Children; 2014.

54. Bergh A-M. Kangaroo mother care implementation workbook. Pretoria: MRC Unit for Maternal and Infant Health Care Strategies; 2002. Available: https://www.healthynewbornnetwork.org/resource/implementation-workbook-for-kangaroo-mother-care. Accessed: 19 September 2021.

55. Community Empowerment Lab. Kangaroo mother care: a hug can save a life. 2016. Available: https://assets.website-files.com/5ef0d72dc95490356f7ad90b/5f0d8d0789c8d58bce4fabcb_Smaply-Case-KMC.pdf. Accessed: 17 November 2021.

56. Gilson L, Schneider H. The impact of free maternal health care in South Africa. In: Berer M, Sundari Ravindran T, editors. Safe motherhood initiatives: critical issues Oxford: Blackwell Science, for Reproductive Health Matters; 4 December 2006. Available: https://www.gov.uk/research-for-development-outputs/the-impact-of-free-maternal-health-care-in-south-africa#contents. Accessed: 3 March 2023.

57. Vani SN. Role of Kangaroo Mother Care Foundation of India in promotion of KMC (unpublished prsentation). XII KMC International Conference, Bogotá, Colombia, 14–17 November 2018. (Updated 2021).

58. Social Finance. Cameroon Kangaroo Mother Care. No date. Available: https://www.socialfinance.org.uk/projects/cameroon-kangaroo-mother-care. Accessed: 29 September 2021.

59. Silver K. The Kangaroo Mother Care Development Impact Bond: Towards worldwide dissemination of KMC (presentation). XII International Conference on Kangaroo Mother Care; Bogotá, Colombia. 2018. Available: https://fundacioncanguro.co/wp-content/uploads/2018/11/KMC-DIB-Karlee-Silver.pdf. Accessed: 20 July 2021.

60. Savell L, Eddleston C. Cameroon KMC Development Impact Bond, 2018-2021. End of Programme Report. September 2021. Available: https://www.socialfinance.org.uk/resources/publications/cameroon-kangaroo-mother-care-development-impact-bond-2018-2021-end-programme. Accessed 29 September 2021.

61. Bergh A-M, Sayinzoga F, Mukarugwiro B, Zoungrana J, Abayisenga G, Karera C, et al. Evaluation of kangaroo mother care services in Rwanda May 2012. Report. Ministry of Health; 2012. Available: https://www.healthynewbornnetwork.org/hnn-content/uploads/12-Rwanda-2012-KMC-review-report.pdf. Accessed: 18 August 2021.

62. Aliganyira P, Kerber K, Davy K, Gamache N, Sengendo NH, Bergh A-M. Helping small babies survive: an evaluation of facility-based Kangaroo Mother Care implementation progress in Uganda. Pan Afr Med J 2014;19:37.

63. International Joint Statement. International Policy Statement for Universal Use of Kangaroo Mother Care for Preterm and Low Birthweight Infants. Commitment to Action from Professional Health Associations; endorsed by the American Academy of Pediatrics (AAP), Council of International Neonatal Nurses (COINN), the International Council of Nurses (ICN), American College of Obstetricians and Gynecologists (ACOG), the International Federation of Gynecology and Obstetrics (FIGO), American College of Nurse-Midwives (ACNM), International Pediatric Association (IPA) and the International Confederation of Midwives (ICM). 17 November 2016. Available: http://ipa-world.org/uploadedbyfck/Professional%20Health%20Care%20Association%20Joint%20statements.pdf. Accessed: 19 November 2016.

64. Stefani G, Skopec M, Battersby C, Harris M. Why is Kangaroo Mother Care not yet scaled in the UK? A systematic review and realist synthesis of a frugal innovation for newborn care. BMJ Innovations. 2022;8(1):9–20.

65. United Nations Development Programme (UNDP). Understanding capacity development 2021. Available: https://www.undp-capacitydevelopment-health.org/en/capacities/. Accessed: 8 October 2021 [

66. Guenther T, Moxon S, Valsangkar B, Wetzel G, Ruiz J, Kerber K, et al. Consensus-based approach to develop a measurement framework and identify a core set of indicators to track implementation and progress towards effective coverage of facility-based Kangaroo Mother Care. J Glob Health. 2017;7(2):020801.

67. Mhango P, Chipeta E, Muula AS, Robb-McCord J, White P, Litch JA, et al. Implementing the Family-Led Care model for preterm and low birth weight newborns in Malawi: Experience of healthcare workers. Afr J Prim Health Care Fam Med. 2020;12(1):a2266.

68. Estifanos AS, Haile Mariam D, Fikre A, Kote M, Tariku A, Chan GJ. Implementation science to design, test and scale up effective Kangaroo Mother Care in Oromia region, Ethiopia. Acta Paediatr. 2022 (online ahead of print).

69. Chan GJ, Labar AS, Wall S, Atun R. Kangaroo mother care: a systematic review of barriers and enablers. Bull World Health Organ. 2016;94(2):130–41.

70. Mathias C, Mianda S, Ohdihambo J, Hlongwa M, Singo-Chipofya A, Ginindza T. Facilitating factors and barriers to kangaroo mother care utilisation in low- and middle-income countries: A scoping review. Afr J Prm Health Care Fam Med. 2021;13(1):a2856.

71. Guenther T, Nsona H, Makuluni R, Chisema M, Jenda G, Chimbalanga E, et al. Home visits by community health workers for pregnant mothers and newborns: coverage plateau in Malawi. J Glob Health. 2019;9(1):010808.

72. Mazumder S, Taneja S, Dube B, Bhatia K, Ghosh R, Shekhar M, et al. Effect of community-initiated kangaroo mother care on survival of infants with low birthweight: a randomised controlled trial. Lancet. 2019;394(10210):1724–36.

73. Spicer N, Hamza YA, Berhanu D, Gautham M, Schellenberg J, Tadesse F, et al. ‘The development sector is a graveyard of pilot projects!’ Six critical actions for externally funded implementers to foster scale-up of maternal and newborn health innovations in low and middle-income countries. Glob Health. 2018;14(1):74.

74. Bergh A-M, de Graft-Johnson J, Khadka N, Om’Iniabohs A, Udani R, Pratomo H, et al. The three waves in implementation of facility-based kangaroo mother care: a multi-country case study from Asia. BMC Int Health Hum Rights 2016;16:4.

75. Bergh A-M, Arsalo I, Malan A, Pattinson R, Patrick M, Phillips N. Measuring implementation progress in kangaroo mother care. Acta Paediatr. 2005;94:1102–8.

76. Belizán M, Bergh A-M, Cilliers C, Pattinson RC, Voce A, for the Synergy Group. Stages of change: A qualitative study on the implementation of a perinatal audit programme in South Africa. BMC Health Serv Res. 2011;11:243.

77. World Health Organization, United Nations Children’s Fund. Every Newborn: an action plan to end preventable deaths. Geneva: World Health Organization; 2014.

78. Hodgins S, Quissell K. Scale-up as if impact mattered: Learning and adaptation as the essential (often missing) ingredient (SNL working paper). December 2016. Available: https://www.healthynewbornnetwork.org/hnn-content/uploads/Empty-Scale-up-Meeting-Report.pdf. Accessed: 28 June 2018.

79. Government of India. Kangaroo Mother Care & Optimal Feeding of Low Birth Weight Infants: Operational guidelines for programme managers & service provider. New Delhi: Ministry of Health and Family Welfare, Child Health Division; 2014. Avalable: https://www.nhm.gov.in/images/pdf/programmes/child-health/guidelines/Operational_Guidelines-KMC_&_Optimal_feeding_of_Low_Birth_Weight_Infants.pdf. Accessed: 21 July 2021.

80. Government of Kenya, Ministry of Health. Kangaroo Mother Care: Clinical Implementation Guidelines. 2016. Available: http://familyhealth.go.ke/wp-content/uploads/2018/02/KANGAROOMOTHERCAREIMPLEMENTATIONGUIDELINES20161.pdf. Accessed: 11 June 2022.

81. Republic of Liberia, Ministry of Health and Social Welfare. Kangaroo Mother Care Guidelines (1^st^ ed). 2013.

82. Western Cape, Kangaroo Mother Care Provinical Task Team. Kangaroo mother care (KMC) policy and guidelines for the Western Cape Province. Cape Town: Western Cape Department of Health; 2003. Available: https://www.westerncape.gov.za/Text/2003/kangaroo_mother_care_policy_guidelines.pdf. Accessed: 20 October 2007.

83. Bergh A-M. “KMC is more than an injection”: Facility-based experiences in selected countries (unpublished presentation). KMC Acceleration Convening, Istanbul, Turkey, 21–22 October 2013.

84. Kachule E. Care of small and early infants. Policy Guidance, Case Studies and Implementation Priorities. Malawi (slides 23-42) (unpublished). Meeting Slides KMC Acceleration Partnership Community of Practice Meeting Blantyre, Malawi, 24th-26th October 2017.

85. Pratomo H. Review of kangaroo mother care in Indonesia, 1997-2014. Unpublished report for Save the Children. 2014.

86. Charpak N, Villegas J, Kangaroo Foundation. Mapping exercise of Kangaroo Mother Care in countries: the example of Colombia (presentation). XI Workshop and Congress of the International Network on Kangaroo Mother Care, Trieste, Italy, 14–17 November 2016. Available: https://fundacioncanguro.co/wp-content/uploads/2018/01/04_charpak_villegas_mapping.pdf. Accessed: 23 July 2021.

87. De Leon-Mendoza S. Kangaroo mother care in the Philippines: assessment of level of implementation. Unpublished research report. December 2013.

88. KMC Acceleration Partnership, Community of Practice. 1st Africa Regional Meeting, 15-17 December 2016, Kigali, Rwanda. Unpublished meeting summary. 2016.

89. KMC Acceleration Partnership, Community of Practice. 1st Asia Regional Meeting, 10-12 December 2016, Dhaka, Bangladesh. Unpublished meeting summary. 2016.

90. KMC Acceleration Partnership, Community of Practice. Meeting, Blantyre, Malawi 24th - 26th October 2017. Unpublished meeting report by Marcella Torres. 2017.

91. KMC Acceleration Partnership, Community of Practice. Africa KAP Supplemental Report, December 15-17, Kigali Rwanda. Unpublished slide presentations of participants. 2016.

92. KMC Acceleration Partnership, Community of Practice. Asia KAP Supplemental Report, December 10-12, Dhaka Bangladesh. Unpublished slide presentations of participants. 2016.

93. KMC Acceleration Partnership, Community of Practice. Meeting Blantyre, Malawi 24th - 26th October 2017. Unpublished slide presentations of participants. 2017.

94. Asia-Oceania Kangaroo Mother Care Network (A-OKNet). Mission-Vision Statement. Unpublished document. 2018.

95. Asia-Oceania Kangaroo Mother Care Network (A-OKNet). Statement of commitment. Unpublished document. 2018.

96. Charpak N. The Kangaroo Mother Care adventure: science and tenderness (unpublished presentation). KMC Acceleration Convening, Istanbul, Turkey, 21–22 October 2013.

97. Charpak N. Lessons learned in implementing kangaroo mother care in Latin American and Caribbean countries through a regional network (presentation). IX International Conference on Kangaroo Mother Care, Ahmedabad, India, 22–25 November 2012. Available: https://fundacioncanguro.co/wp-content/uploads/2017/10/9ICKMC-OP-49-Nathalie-Charpak.pdf. Accessed: 23 July 2021.

98. Save the Children. The Regional Learning Network: A Model for Improving Maternal and Newborn Health Care Outcomes in Uganda. 2018. Avalable: https://resourcecentre.savethechildren.net/node/18078/pdf/uganda-rln-2020-final.pdf. Accessed: 12 November 2021.

99. Ediamu T, Pirio P, Beyeza-Kashesya J, Ongom S. The Regional Learning Network: A model for improving maternal and newborn health care outcomes in Uganda (Poster). 2018. Available: https://resourcecentre.savethechildren.net/library/regional-learning-network-model-improving-maternal-and-newborn-health-care-outcomes-uganda. Accessed: 12 November 2021.

100. Ediamu T, Pirio P. UGANDA: Establishing a Regional Learning Network to Improve Quality of Care for Mothers and Newborns (presentation). XI Workshop and Congress of the International Network on Kangaroo Mother Care, Trieste, Italy, 14–17 November 2016. Available: https://fundacioncanguro.co/wp-content/uploads/2018/01/19_ediamu_uganda.pdf. Accessed: 12 November 2021.

101. Kinney M, Corber E. “A lot has changed…”: Reflections on the Impact of the Regional Learning Network initiative in Uganda (blog). 28 April 2020. Available: https://www.healthynewbornnetwork.org/blog/a-lot-has-changed-reflections-on-the-impact-of-the-regional-learning-network-initiative-in-uganda/. Accessed: 12 November 2021.

102. Save the Children. From Invisibility to Value: Improving quality of care for small and sick newborns (presentation). IHI Africa Forum on Quality and Safety in Healthcare, Durban, South Africa, February 2018. Available: https://resourcecentre.savethechildren.net/library/invisibility-value-improving-quality-small-and-sick-newborns. Accessed: 27 July 2021.

103. Maternal and Child Survival Program (MCSP)/USAID. Kangaroo Mother Care in Malawi. 2017. Available: https://www.mcsprogram.org/wp-content/uploads/dlm_uploads/2019/01/Malawi-KAP-Summary-Sheet.pdf: Accessed: 23 October 2021.

104. Longwe M. Khanda ndi Mphatso Campaign Report for Save the Children. June 2016. Available: https://www.healthynewbornnetwork.org/resource/khanda-ndi-mphatso-sbcc-campaign-report/. Accessed: 23 October 2021.

105. Banda G. Khanda Ndi Mphatso (A Baby is a Gift): Evaluation of a pilot SBCC campaign to shift social norms & care practices for preterm and low birthweight babies (presentation). Save the Children International Malawi SNL Legacy Webinar, 1 October 2020. Available: https://www.healthynewbornnetwork.org/hnn-content/uploads/Gedesi-B_SBCC-presentation_Malawi_Oct-6-SNL-Legacy_v2.pdf. Accessed: 26 October 2021.

106. Save the Children. “Khanda Ndi Mphatso”: Sustaining and expanding activities to shift social norms and care practices for preterm and low birthweight babies (brief). April 2020. Available: https://resourcecentre.savethechildren.net/node/18066/pdf/sbcc-brief_web.pdf. Accessed: 27 July 2021.

107. Robb-McCord J, Greensides D, Kamanga E, Litch J. Formative assessment of community, family and health care provider knowledge, attitudes, beliefs and practices regarding preterm and low birth weight newborns in Balaka district, Malawi Every Preemie— SCALE; 2017. Available: https://www.everypreemie.org/wp-content/uploads/2016/11/JUNE-2017-Malawi-Formative-Assessment-Balaka-District-FINAL.pdf. Accessed: 25 July 2019.

108. Bergh A-M, Hoque D, Udani RH, Rao S, Puri A, Pratomo H, et al. The implementation and scale up of facility-based kangaroo mother care in five Asian countries. Unpublished research report for Save the Children and USAID/MCHIP. September 2014.

109. Bergh A-M, Manu R, Davy K, Van Rooyen E, Quansah-Asare G, Awoonor-Williams JK, et al. Translating research findings into practice - the implementation of kangaroo mother care in Ghana. Implement Sci. 2012;7:75.

110. Bergh A-M, Kerber K, Abwao S, de-Graft Johnson J, Aliganyira P, Davy K, et al. Implementing facility-based kangaroo mother care services: lessons from a multi-country study in Africa. BMC Health Serv Res. 2014;4:293

111. Miller-Petrie MK, Mazia G, Serpa M, Pooley B, Marshall M, Meléndez C, et al. Building alliances for improving newborn health in Latin America and the Caribbean. Revista panamericana de salud publica = Pan Am J Public Health. 2014;36(1):44–9.

112. Bergh A-M, Allanson E, Pattinson R. What is needed for taking emergency obstetric and neonatal programmes to scale? Best Pract Res Clin Obstet Gynaecol. 2015;29(8):1017–27.

113. Kululanga LI, Sundby J, Malata A, Chirwa E. Male involvement in maternity health care in Malawi. Afr J Reprod Health. 2012;16(1):145–57.

114. Manda-Taylor L, Mwale D, Phiri T, Walsh A, Matthews A, Brugha R, et al. Changing times? Gender roles and relationships in maternal, newborn and child health in Malawi. BMC Pregnancy Childbirth. 2017;17(1):321.

115. Social Finance. Kangaroo mother care (KMC) DIB: Lessons from delivery for scale. March 2021. Available: https://www.socialfinance.org.uk/sites/default/files/publications/cameroon_kmc_dib_-_lessons_from_delivery_for_scale.pdf. Accessed: 29 September 2021.

116. Bergh A-M. Implementation and evaluation of kangaroo mother care in ten hospitals in Indonesia, Unpublished report to USAID/Indonesia Health Services Program/JSI, Inc.; 2011.

117. Gontijo TL, Meireles AL, Malta DC, Proietti FA, Xavier CC. Evaluation of implementation of humanized care to low weight newborns - the Kangaroo Method. J Pediatr (Rio J). 2010;86(1):33–9.

118. Welch PR, Kavle JA, Bwanali F, Nyambo K, Khadka N. A Health System Bottleneck Analysis of Care and Feeding of Small and Sick Newborns in Malawi: Findings and Considerations for Nutrition-Newborn Integration. Final Report. USAID/MCSP; 2015. Available: https://www.mcsprogram.org/resource/a-health-system-bottleneck-analysis-of-care-and-feeding-of-small-and-sick-newborns-in-malawi-findings-and-considerations-for-nutrition-newborn-integration/. Accessed 29 September 2021.

119. Logie DE, Rowson M, Ndagije F. Innovations in Rwanda’s health system: looking to the future. Lancet. 2008;372(9634):256-61.

120. Mehjabeen S, Matin M, Gupta RD, Sutradhar I, Mazumder Y, Kim M, et al. Fidelity of kangaroo mother care services in the public health facilities in Bangladesh: a cross-sectional mixed-method study. Implement Sci Communications. 2021;2(1):115.

121. Colombia Ministry of Health and Social Protection (MINSALUD). Actualización de los Lineamientos Técnicos para la implementación de Programas Madre Canguro en Colombia, con énfasis en la nutrición del neonato prematuro [Update of the Technical Guidelines for the Implementation of Kangaroo Mother Programs in Colombia with emphasis on nutrition for premature or low birth weight infants]. Bogotá, Colombia: MINSALUD; November 2017. Available: https://www.minsalud.gov.co/sites/rid/Lists/BibliotecaDigital/RIDE/DE/implementacion-programa-canguro.pdf and http://fundacioncanguro.co/wp-content/uploads/2019/06/Update-of-the-Technical-Guidelines-for-the-Implementation-of-Kangaroo-Mother-Programs-in-Colombia-P.pdf. Accessed: 20 July 2021.

122. Bergh A-M. Trip report and action points. Unpublished report to UNICEF on trip to Chad. 2017.

123. Chavula K, Likomwa D, Valsangkar B, Luhanga R, Chimtembo L, Dube Q, et al. Readiness of hospitals to provide Kangaroo Mother Care (KMC) and documentation of KMC service delivery: Analysis of Malawi 2014 Emergency Obstetric and Newborn Care (EmONC) survey data. J Glob Health. 2017;7(2):020802.

124. International Centre for Diarrhoeal Diseases Research, Bangladesh. To explore the feasibility of introduction and operational challenges of implem entation of Kangaroo Mother Care (KMC) services in selected first level referral facilities of Kushtia, Bangladesh. 2017. Available: https://www.healthynewbornnetwork.org/hnn-content/uploads/OR-KMC_Baseline_Report_20150808.pdf. Accessed: 24 July 2021.

125. Ahmed S, Alam S, Mannan I, Khan M, George J. Overcoming health systems bottlenecks in implementing Kangaroo Mother Care at district and sub-district level health facilities in Bangladesh (poster). XI Workshop and Congress of the International Network on Kangaroo Mother Care, Trieste, Italy, 14–17 November 2016. Available: https://fundacioncanguro.co/wp-content/uploads/2018/01/06_Marufa-Aziz-Khan_poster.pdf. Accessed: 23 July 2021.

126. Manji K. Situation analysis of newborn health in Tanzania: Current situation, existing plans and strategic next steps for newborn health. Dar es Salaam: Ministry of Health and Social Welfare, Save the Children; 2009. Available: https://resourcecentre.savethechildren.net/pdf/6390.pdf/. Accessed: 21 June 2021.

127. Bar-Zeev N, Gladstone M, Kungwimba E, Ngwria B, Dube Q, Bar-Zeev S. Situational analysis of newborn health in Malawi 2013. Blantyre: Reproductive Health Unit, Malawi Ministry of Health; Save the Children; Support for Service Delivery Integration (SSDI); UNICEF; World Health Organization; 2013. Available: https://www.scribd.com/document/300019739/Malawi-s-Newborns-Report-2013. Accessed: 20 June 2021.

128. Government of Bangladesh, Ministry of Health and Family Welfare. BANGLADESH National Newborn Health Situation Analysis Report. 2014. Available: https://www.exemplars.health/-/media/files/egh/resources/underfive-mortality/bangladesh/unicef_bangladesh-national-newborn-health-situation-analysis-report.pdf?la=en. Accessed: 24 July 2021.

129. Hoque DME. Situation analysis report on the introduction and feasibility of the expansion of kangaroo mother care (KMC) in Bangladesh. Unpublished report for Save the Children and USAID/MCHIP. December 2013.

130. Engmann C. The Case for KMC Acceleration (unpublished presentation). KMC Acceleration Convening; Istanbul, Turkey, 21–22 October 2013.

131. Cattaneo A, Amani A, Charpak N, De Leon-Mendoza S, Moxon S, Nimbalkar S, et al. Report on an international workshop on kangaroo mother care: lessons learned and a vision for the future. BMC Pregnancy Childbirth. 2018;18(1):170.

132. Salim N, Shabani J, Peven K, Rahman QS-U, Kc A, Shamba D, et al. Kangaroo mother care: EN-BIRTH multi-country validation study. BMC Pregnancy Childbirth. 2021;21(Suppl 1):231.

133. Day LT, Sadeq-ur Rahman Q, Ehsanur Rahman A, Salim N, Kc A, Ruysen H, et al. Assessment of the validity of the measurement of newborn and maternal health-care coverage in hospitals (EN-BIRTH): an observational study. Lancet Glob Health. 2021;9(3):e267–e79.

134. Bangladesh Ministry of Health and Family Welfare, Save the Children. Brief 6: Implementing the Comprehensive Newborn Care Package: Integration and Use of Data in Routine Health Information System. November 2017. Available: https://www.healthynewbornnetwork.org/hnn-content/uploads/06.HIS_.pdf. Accessed: 25 July 2021.

135. Charpak N, Angel MI, Banker D, Bergh AM, María Bertolotto A, De Leon-Mendoza S, et al. Strategies discussed at the XIIth international conference on Kangaroo mother care for implementation on a countrywide scale. Acta Paediatr. 2020;109(11):2278–86.

136. Pratomo H, Uhudiyah U, Poernomo Sigit Sidi I, Rustina Y, Suradi R, Bergh A-M, et al. Supporting factors and barriers in implementing kangaroo mother care in Indonesia. Paediatr Indones. 2012;52(1):43–50.

137. Weldearegay HG, Medhanyie AA, Abrha MW, Tadesse L, Tekle E, Yakob B, et al. Quality of Kangaroo Mother Care services in Ethiopia: Implications for policy and practice. PloS One. 2019;14(11):e0225258.

138. Nimbalkar S, Sadhwani N. Implementation of Kangaroo Mother Care - Challenges and Solutions. Indian Pediatr. 2019;56(9):725–9.

139. Welch P, Charpak N. An Assessment of the Status of Kangaroo Mother Care in the Dominican Republic: Findings and Considerations for Sustainability. Final Report. USAID/MCSP; 2019. Available: https://www.healthynewbornnetwork.org/hnn-content/uploads/MCSP-DR-KMCAssessmentReport.pdf. Accessed: 13 November 2021.

140. Maternal and Child Survival Program (MCSP)/USAID, Save the Children. Tracking implementation progress of Kangaroo Mother Care: Overview of results of a multi-country evaluation. 2014. Available: https://www.mchip.net/sites/default/files/Tracking%20Implementation%20Progress%20for%20KMC.pdf. Accessed: 23 July 2021.

141. Bergh A-M, Sylla M, Traore I, Diall Bengaly H, Kante M, Kaba D. Evaluation of kangaroo mother care services in Mali. Unpublished report. Washington, DC: Save the Children; 2012. Available: https://www.mchip.net/sites/default/files/Mali%20KMC%20Report.pdf. Accessed: 27 April 2021.

142. Working group. Group work on enablers and barriers, 14 November 2016: India and Brazil. 2016. Available: https://fundacioncanguro.co/documentos/?lang=en# (XI International Conference on KMC – Trieste, Italy 2016; Workshop; WG14indiabrazil_report). Accessed: 8 April 2022.

143. Bangladesh Ministry of Health and Family Welfare. National Newborn Health Program implementation toolkit (2nd ed). June 2019. Available: https://www.researchgate.net/publication/343046423_National_Newborn_Health_Program_Implementation_Toolkit. Accessed: 21 June 2021

144. Bergh A-M, Van Rooyen E, Pattinson RC. Scaling up kangaroo mother care in South Africa: ‘on-site’ versus ‘off-site’ educational facilitation. Human Resources Health. 2008;6:13.

145. Bergh A-M, Charpak N, Enzeonodo A, Udani R, Van Rooyen E, for the KMC Education and Training Working Group. Education and training in the implementation of kangaroo mother care. S Afr J Child Health. 2012;6(2):38–45.

146. Foote N, Tamburlini G. Accelerating the effective scaling of Kangaroo Mother Care and related interventions: lessons from 20 years of experience. Early Childhood Matters (Bernard van Leer Foundation). 2017. Available: https://bernardvanleer.org/ecm-article/2017/accelerating-effective-scaling-kangaroo-mother-care-related-interventions-lessons-20-years-experience/. Accessed: 10 October 2021.

147. Bangladesh Ministry of Health and Family Welfare, Children St. Brief 3: Facility readiness and initiation of kangaroo mother care. November 2017. Avalable: https://www.healthynewbornnetwork.org/resource/facility-readiness-and-initiation-of-kangaroo-mother-care/. Accessed: 27 July 2021.

148. Maternal and Child Survival Program (MCSP)/USAID. Kangaroo Mother Care in Bangladesh. 2017. Available: https://www.mcsprogram.org/wp-content/uploads/dlm_uploads/2019/01/Bangladesh-KAP-Summary-Sheet.pdf. Accessed: 23 October 2021.

149. Pattinson RC, Arsalo I, Bergh A-M, Malan AF, Patrick M, Phillips N. Implementation of kangaroo mother care: a randomized trial of two outreach strategies. Acta Paediatr. 2005;97(4):924–7.

150. Department of Health South Africa. Essential Newborn Care Quality Improvement Toolkit. 2013. Available: http://www.lincare.co.za/?page_id=1143. Accessed: 18 November 2021.

