## Supplemental file 1 for "Programmatic implementation of kangaroo mother care: a systematic synthesis of grey literature"

### Primary screening route

The figure below is a graphic depiction of the primary screening route. The 753 documents subjected to the first screening were not recorded on a data extraction sheet.

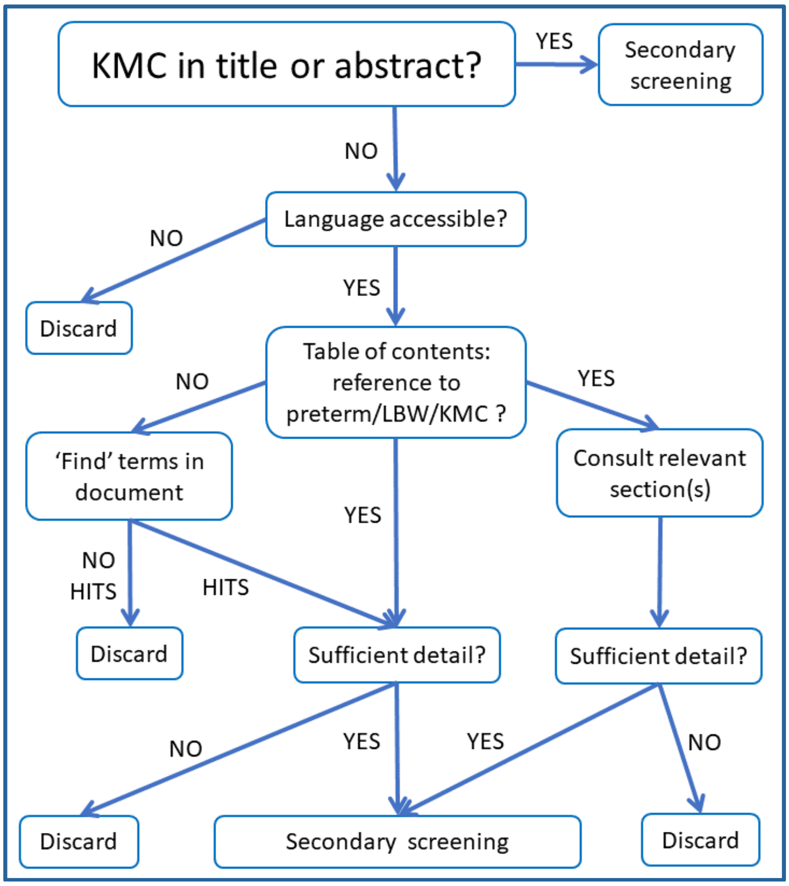


The primary screening included the following:

Does the title (or abstract, where applicable) contain terms like "kangaroo mother care", "kangaroo care" or "skin-to-skin care"? YES / NO

If YES: **Go directly to the secondary screening**

If NO: Do further screening

↓

- Language that the consultant understands? YES / NO
  - If YES: Continue below
  - If NO: Is it possible to do a Google translation of the title and table of contents? YES / NO
    - If YES: Do translation of title, table of contents and search terms and continue below
    - If NO: Consult with WHO for assistance OR eliminate from further screening and analysis
- Scan the table of contents: Any reference to preterm / low birthweight or KMC? YES / NO
  - If YES: Consult the relevant section(s) of the document
  - If NO: Do a word search for the following terms: "preterm", "prematur*", "low birth weight", "low birthweight", "LBW", "kangaroo", "KMC"

- Apply screening questions for the total document:
    - Did the search with the above search terms yield any hits? YES / NO
        - If NO: Discard document
        - If YES: KMC discussed apart from one-sentence mentions as method of care for small babies? YES / NO
            - If NO: Discard document
            - **If YES: Continue with secondary screening**
