## Supplemental file 2 for "Programmatic implementation of kangaroo mother care: a systematic synthesis of grey literature"

**Supplemental file 2 DATABASE OF WEBSITES AND REPOSITORIES**

For further exploration (highlighted)

| Date | Name of website | URL | Search terms | # hits | Saved | File name | Comments | Identified for secondary screening |
|---|---|---|---|---|---|---|---|---|
| 2021/06/24 | ClinicalTrials.gov | https://www.clinica | kangaroo mother care AND preterm birth | 38 | Y | 2021-06-04 ClinicalTrial | None of the titles indicate relevance<br>The Norwegian Family Centered Care Study compared two unit set-ups: **family centered vs open-bay unit** - no outcome related to coverage | |
| 2021/06/25 | Sigma repository | https://sigma.nurs | TITLE: (kangaroo AND | 1701 | Y | 2021-06-25 Sigma repos | Checked the first 100 titles (sorted according to relevance): only 8 have KMC or KC in title | |
| 2021/06/25 | NLM Bookshelf | https://www.ncbi.n | TITLE: (kangaroo mothe | 0 | N | N/A | No books with KMC in title | |
| 2021/06/25 | OAIster | https://oaister.wor | TITLE: kangaroo mothrs car | 4 | Y | 2021-06-25 OAIster Res | Filter: Theses and dissertations (2 English, 1 Italian & 1 Portuguese)<br>None of titles indicate relevance | |
| 2021/06/25 | The New York Academy of Medicine - Grey Literature | http://www.greylit. | kangaroo mother care | 0 | N | - | | |
| 2021/06/25 | African Digital Health Library | http://adhlui.com.u | - | 0 | Y | 2021-06-25 African Digit | OpenDOAR - Preterm not a tab to click - only "pregnant women" and "antenatal care" | |
| 2021/06/25 | ACHS, Theses and Capstone Projects | https://www.achs. | - | 0 | N | - | OpenDOAR - Link did not work | |
| 2021/06/25 | All Ireland Public Health Repository | http://repository.p | kangaroo mother care AND preterm birth | 0 | Y | 2021-06-25 All Ireland P | OpenDOAR - Way the sources is organised does not at a first glance give an indication of finding something useful | |
| 2021/05/25 | Advocate Aurora Health Insitutional Repository | https://institutiona | kangaroo mother care OR kangaroo care | 0 | Y | 2021-06-25 Advocate A | OpenDOAR - I hit not relevant - downlaoded source | |
| 2021/06/25 | BRAC Univeristy Insitutional Repository | http://dspace.brac | kangaroo mother care OR kangaroo care | 12 | Y | 2021-06-25 BRAC Univ | OpenDOAR - None of the documents on the list relevant to our focus - 6 hits = annual reports<br>2 documents related to the **Quasem et al 2002/3 study (community based implementation of KMC)**<br>Downloaded Brenner's article on signal functions - KMC not mentioned, only a | |
| 2021/06/25 | BiblioDigital SIDRE | https://v2.sherpa.a | - | 0 | N | - | OpenDOAR - Link did not work (Colombian databse; Spanish interface) | |
| 2021/06/25 | Biblioteca Virtual em Saúde - Minisétrio da Saúde (official MoH site) | https://www.gov.b | canguru | 958 | Y | 2021-06-25 Brazil MoH | OpenDOAR - Browsed through first 6 pages (± 150 hits) and did some google translations - potential titles to access marked in yellow on the saved files - PORTUGUESE | |
| 2021/06/25 | Brazilian Hospital Services Company | https://www.gov.b | canguru (Got on through the Brazilian MoH) | 1 | Y | Brazil Port 00814-18 - D | Irrelevant dissertation | |
| 2021/06/25 | CARE - CCLHD Archive and Research E-Library (CCLHD) | http://centralcoast | kangaroo mother care OR kangaroo care | 0 | Y | 2021-06-25 CCLHD Arc | OpenDOAR - No references | |
| 2021/06/25 | Electronic Theses and Dissertations of The Tamil Nadu Dr. M.G.R. Medical University | http://repository-tn | Items where Subject is "MEDICAL > Neonatology" | 19 | Y | 2021-06-25 EPrints@Ta | OpenDOAR - No relevant references | |

**Supplemental file 2 DATABASE OF WEBSITES AND REPOSITORIES**

For further exploration

**Identified for secondary screening**

| Date | Name of website | URL | Search terms | # hits | Saved | File name | Comments |
|---|---|---|---|---|---|---|---|
| 2021/06/25 | Health Sciences Research Commons (George Washington Schools of Medicine and Health Sciences) | https://hsrc.himme | kangaroo mother care OR kangaroo care | 6 | Y | 2021-06-25 Health Scie | OpenDOAR 6 publications (articles) listed, i.a. systematic reviews by Chan et al and Smith et al - they will be picked up by the systematic review if relevant |
| 2021/06/25 | Humboldt Digital Scholar | https://humboldt-d | kangaroo mother care OR kangaroo care (Title) | 0 | N | - | OpenDOAR - No references |
| 2021/06/25 | Indian Academy of Sciences: Publications of Fellows | http://repository.ia | kangaroo mother care OR kangaroo care | 0 | N | - | OpenDOAR - No references |
| 2021/06/26 | icddr,b (Bangladesh) Knowledge Repository | http://dspace.icdd | kangaroo mother care OR kangaroo care | 0 | N | - | OpenDOAR - No references |
| 2021/06/26 | Knowledge Repository Open Network (India) | http://dspaces.uol | - | 0 | N | - | OpenDOAR - Link did not work |
| 2021/06/26 | LSHTM Research Online | http://researchonl | kangaroo mother care kangaroo care | 10 | Y | 2021-06-25 LSHTM Res | Open DOAR 9 references = published KMC articles; 1 = thesis by article by Medvedev Search with KC yielded the same 10 hits |
| 2021/06/26 | Norwegian Institute of Public Health Open Repository | https://fhi.brage.u | kangaroo mother care OR kangaroo care | 8 | Y | 2021-06-26 Norwegian I | Open DOAR - 3 references in NORWEGIAN - rest = published articles (Yoshida et al & Ames et al - only mention of KMC |
| 2021/06/26 | Open Knowledge Repository (World Bank) | https://openknowl | kangaroo mother care kangaroo care | 0 | N | 2021-06-26 Open Know | OpenDOAR - No references |
| 2021/06/26 | OpenPub (CINECA IRIS Institutional Research Information System) | https://openpub.fr | kangaroo mother care kangaroo care | 0 | N | - | OpenDOAR - No references |
| 2021/06/26 | PubMed Central | http://www.ncbi.nl | (kangaroo mother care[T | 841 | N | - | OpenDOAR - publications articles and books - will be covered by the systematic review |
| 2021/06/26 | IRIS PubliCat (Itallian)t | https://publicatt.ur | kangaroo mother care | 6 | Y | 2021-06-26 PubliCatt (E | OpenDOAR - None relevant |
| 2021/06/16 | Repository of the Iran University of Medical Sciences, Tehran | | kangaroo mother care kangaroo care | 0 | Y | 2021-06-26 Iran Univers | OpenDOAR - No references |
| 2021/06/26 | SciELO Public Health (Brazil) | http://www.scielos | kangaroo method | 4 | Y | 2021-06-26 SciELO Pub | OpenDOAR - 4 references in PORTUGUESE |
| | | | kangaroo care | 9 | Y | | 9 references = combining the references from the other 2 searches |
| | | | kangaroo mother care | 5 | Y | | 5 references - 3 PORTUGUESE; 1 SPANISH, 1 English |

**Supplemental file 2 DATABASE OF WEBSITES AND REPOSITORIES**

For further exploration

| Date | Name of website | URL | Search terms | # hits | Saved | File name | Comments | Identified for secondary screening |
|---|---|---|---|---|---|---|---|---|
| 2021/06/26 | Argentina: Scientific Electronic Library Online (Scielo) | http://www.scielo. | kangaroo mother care [ | 0 | Y | 2021-06-26 Scielo Arge | OpenDOAR - No references | |
| 2021/06/26 | Brazil: Scientific Electronic Library Online | http://www.scielo. | kangaroo care (title) | 37 | Y | 2021-06-26 SciELO Bra | OPenDOAR - References and Eng abstracts downloaded<br>**All references (with an English abstract) downloaded (n=37) = mostly journal articles** | 37 |
| 2021/06/26 | Chile: Scientific Electronic Library Online | http://www.scielo. | kangaroo mother care [ | 0 | N | - | OpenDOAR - No references | |
| 2021/06/26 | Colombia: Scientific Electronic Library Online | http://www.scielo. | kangaroo mother care [ | 0 | N | - | OpenDOAR - No references | |
| 2021/06/26 | Costa Rica: Scientific Electronic Library Online | http://www.scielo. | kangaroo mother care [ | 0 | N | - | OpenDOAR - No references | |
| 2021/06/26 | Cuba: Scientific Electronic Library Online | http://www.scielo. | kangaroo mother care [ | 0 | N | - | OpenDOAR - No references | |
| 2021/06/26 | Mexico: Scientific Electronic Library Online | http://www.scielo. | - | 0 | N | N/A | Could not access the database | |
| 2021/06/26 | Paraguay: Scientific Electronic Library Online | http://scielo.iics.u | kangaroo mother care [ | 0 | N | - | OpenDOAR - No references | |
| 2021/06/26 | Perú: Scientific Electronic Library Online | http://www.scielo. | kangaroo mother care [ | 0 | N | - | OpenDOAR - No references | |
| 2021/06/26 | Portugal: Scientific Electronic Library Online | http://www.scielo. | kangaroo mother care [ | 0 | N | - | OpenDOAR - No references | |
| 2021/06/26 | Spain: Scientific Electronic Library Online | http://scielo.isciii. | kangaroo mother care [ | 0 | N | - | OpenDOAR - No references | |
| 2021/06/26 | Uruguay: Scientific Electronic Library Online | http://www.scielo. | kangaroo mother care [ | 0 | N | - | OpenDOAR - No references | |
| 2021/06/26 | medRxiv | https://www.medr | kangaroo mother care OR kangarr=oo care OR kangaroo method | 1 | N | - | OpenDOAR - one relevant reference:<br>Disparities in Kangaroo Care for Premature Infants in the Neonatal Intensive Care Unit<br>Edith Brignoni-Pérez, Melissa Scala, Heidi M. Feldman, Virginia A. Marchman, Katherine E. Travis<br>medRxiv 2020.11.09.20224766; doi: https://doi.org/10.1101/2020.11.09.20224766 | |
| 2021/07/07 | ProQuest Dissertations & Theses Global (UP access) | https://www-proqu | Kangaroo Mother Care | 8185 | Y | 2021-07-07 ProQuest D | All hits - has captured all "kangaroo care" combinations - same for basic and advanced search<br>Only 4 theses/dissertations with the full name of "kangaroo mother care in the title<br>18 our of first 100 hits have "kangaroo care" in their title | |

**Supplemental file 2 DATABASE OF WEBSITES AND REPOSITORIES**

For further exploration

| Date | Name of website | URL | Search terms | # hits | Saved | File name | Comments | Identified for secondary screening |
|---|---|---|---|---|---|---|---|---|
| 2021/07/07 | ProQuest Dissertations & Theses Global (UP access) | https://www-proqu | Kangaroo Mother Care | 7631 | Y | 2021-07-07 ProQuest D | Filter: full text | |
| 2021/07/07 | ProQuest Dissertations & Theses Global (UP access) | https://www-proqu | Kangaroo Mother Care | 6483 | Y | 2021-07-07 ProQuest D | Filter Doctoral dissertations only | |
| 2021/07/07 | ProQuest Dissertations & Theses Global (UP access) | https://www-proqu | Kangaroo Mother Care AND scale-up | 109 | Y | 2021-07-08 ProQuest D | All hits - also picks up "kangaroo care" and "kangaroo mother method" | |
| 2021/07/08 | ProQuest Dissertations & Theses Global (UP access) | https://www-proqu | Kangaroo Mother Care AND scale up | 6946 | N | - | All hits - NB spelling matters! (Scale up without a hyphen) | |
| 2021/07/08 | ProQuest Dissertations & Theses Global (UP access) | https://www-proqu | ti(kangaroo mother care) OR ab(kangaroo mother care) | 42 | Y | 2021-07-08 ProQuest D | All hits - marked 10 | |
| 2021/07/08 | ProQuest Dissertations & Theses Global (UP access) | https://www-proqu | ti(kangaroo mother care) OR ab(kangaroo mother care) | 10 | Y | 2021-07-08 ProQuest D | Abstracts included<br>**3 identified for secondary screening - PDFs downloaded** | 3 |
| 2021/07/08 | ProQuest Dissertations & Theses Global (UP access) | https://www-proqu | ti(implementation) OR ab(implementation) | 203,962 | N | - | Only "implementation" | |
| 2021/07/08 | ProQuest Dissertations & Theses Global (UP access) | https://www-proqu | ti(scale-up) OR ab(scale-up) | 3711 | N | - | Only "scale-up" | |
| 2021/07/08 | ProQuest Dissertations & Theses Global (UP access) | https://www-proqu | ti(scale up) OR ab(scale up) | 44,866 | N | - | Only "scale up" | |
| 2021/07/08 | ProQuest Dissertations & Theses Global (UP access) | https://www-proqu | ti(kangaroo mother care | 7 | N | - | Same titles already identified in previous searches | |
| 2021/07/08 | Harvard Kennedy School | https://cse.google | kangaroo mother care | 25 | Y | 2021-07-08 Harvard Ker | University of Illinois Chicago Public Health<br>**4 documents on development impact bonds - earmarked for secondary screening**<br>2 documents on KMC coverage estimates by the Guttmacher Institute forwarded to Nils | 4 |

**Supplemental file 2 DATABASE OF WEBSITES AND REPOSITORIES**

For further exploration

| Date | Name of website | URL | Search terms | # hits | Saved | File name | Comments | Identified for secondary screening |
|---|---|---|---|---|---|---|---|---|
| 2021/07/08 | National Library of Medicine | https://www.nlm.n | kangaroo mother care | 304 | Y | 2021-07-08 National Lib | University of Illinois Chicago Public Health<br>Only first 3 documents pertain ot KMC = changes in MeSH terms | |
| 2021/07/08 | WHO | https://www.who.i | kangaroo mother care | 8 | Y | 2021-07-08 WHO searc | University of Illinois Chicago Public Health<br>**1 document for secondary screening** | 1 |
| 2021/07/08 | MetaLib | https://metalib.gp | kangaroo mother care | 95 | Y | 2021-07-08 MetaLib® - | University of Illinois Chicago Public Health<br>Only first 31 hits relevant - no documents identified | |
| 2021/07/09 | congress.gov | https://www.congr | kangaroo mother care | 0 | Y | 2021-07-09 Congress.g | University of Illinois Chicago Public Health<br>No hits | |
| 2021/07/09 | USA.gov | https://www.usa.g | kangaroo mother care | Unknown | Y | 2021-07-09 USA.gov Se | University of Illinois Chicago Public Health<br>All Pudmed references in this list | |
| 2021/07/11 | Scopus | - | - | - | - | - | Only journal titles | |
| 2021/07/11 | EBSCOhost | UP library | TI kangaroo mother care OR TI kangaroo care OR AB kangaroo mother care OR AB kangaroo care | 1921 | Y | EBSCOhost details of d | Databases iincluded: Academic Search Complete; APA PsycArticles; APA PsycInfo; ERIC; Family and Society Studies Worldwide; Health Source - Consumer Edition; Health Source: Nursing/Academic Edition; MasterFILE Premier; Social Work Abstracts; TOC Premier; Open Dissertations | |
| 2021/07/11 | EBSCOhost | UP library | TI kangaroo mother care OR TI kangaroo care OR AB kangaroo mother care OR AB kangaroo care | 206 | Y | EBSCOhost Health Sou | Databases iincluded: Health Source - Consumer Edition; Health Source: Nursing/Academic Edition<br>Most were papers in academic journals - 9 hits that were in periodicals and one book review - none relevant for our task | |
| 2021/07/11 | Agency for Healthcare Research and Quality | https://effectivehe | kangaroo mother care | 6 | Y | 2021-07-11 AHRQ Sear | Nothing of use - 5 references related to a literature review (CHIPRA) - review downloaded and filed under "Irrelevant sources downloaded" (chipra-119-section-5-lit-review.pdf)<br>1 reference to a White Paper on Global Health Evidence Evaluation Framework - **downloaded for secondary screening** | 1 |
| 2021/07/11 | EBSCO Open Dissertations | | | | | | **SIGN IN ON UP WEBSITE** | |
| 2021/07/11 | BMC Proceedings | https://bmcproc.bi | kangaroo mother care OR kangaroo care OR skin-to-skin | 1 | Y | BMC Proceedings _ Arti | La Trobe University<br>Not relevant | |
| 2021/07/11 | Explore the British Library | http://explore.bl.u | kangaroo mother care | 449 | Y | 2021-07-11 Explore Brit | La Trobe University<br>Searched ALL items - mostly journal titles<br>Revisit later - journal titles have aspects that relate to health system building blocks | |

**Supplemental file 2 DATABASE OF WEBSITES AND REPOSITORIES**

For further exploration

| | Date | Name of website | URL | Search terms | # hits | Saved | File name | Comments | Identified for secondary screening |
|---|---|---|---|---|---|---|---|---|---|
| | 2021/07/11 | NZ Ministry of Health - Grey Matter Newsletter | https://www.health | kangaroo mother care | 170 | N | - | La Trobe University<br>No articles on KMC - seach separates the three words - a lot of breastfeeding (probably with words "mother" and "care") | |
| | 2021/07/11 | Open Grey | http://www.opengr | kangaroo mother care | 1 | N | - | La Trobe University<br>Only a thesis on drug addiction | |
| | 2021/07/12 | Internet Archive (WayBack Machine) | https://archive.org | kangaroo mother care | 0 | Y | 2021-07-12 Internet Arch | Found on Georgetown University (Dahlgren Memorial Library) only tracks number of times a webiste was consulted | |
| | 2021/07/12 | Web of Science | Via UP library | TS=(kangaroo mother care) (exact search) | 1577 | Y | 2021-07-12 WoS TS=(ka | Mostly journal articles - revisit at a later stage | |
| | 2021/07/13 | Slideshare | https://www.slides | kangaroo mother care | 3892 | Y | 2021-07-13 SlideShare ' | Accessed via LinkedIn<br>Revisit later with refined searches | |
| | 2021/07/13 | Slideshare | https://www.slides | kangaroo mother care AND implementation | 890 | N | - | Accessed via LinkedIn<br>Revisit later with refined searches | |
| | 2021/07/13 | Slideshare | https://www.slides | kangaroo mother care AND scale up | 2118 | N | - | Accessed via LinkedIn<br>Revisit later with refined searches | |
| | 2021/07/13 | Slideshare | https://www.slides | kangaroo mother care AND (scale up OR scaling up) | 322 | N | - | Accessed via LinkedIn<br>Revisit later with refined searches | |
| | 2021/07/13 | Slideshare | https://www.slides | kangaroo mother care AND (implementation OR scale up OR scaling up) | 189 | N | - | Accessed via LinkedIn<br>Revisit later with refined searches | |
| | 2021/07/13 | Slideshare | https://www.slides | kangaroo mother care AND coverage | 1056 | N | - | Accessed via LinkedIn<br>Revisit later with refined searches | |
| | 2021/07/13 | Trove (Australia) | https://trove.nla.go | kangaroo mother care | 410 | Y | 2021-07-13 Trove | Accessed via Monash University<br>Revisit later - also magazine article - quite a number on kangaroos and not KMC | |
| | | **WEBSITES IN KC BIB:** | | | | | | | |
| 1 | 2021/07/14 | United States Institute for Kar | www.kangaroocareusa.org | | **NO RELEVANT RESOURCES** | | | KC BIB: 1.The United States Institute for Kangaroo Care is developing their website at www.kangaroocareusa.org and has two emails in place: Email them for information and they check for emails at least 3 times/ week. This a resource center, telling you where to get chairs, videos, and making arrangements for speakers and customized speaking needs as well as consultations and instruction on the floors and units where KMC is found. | |
| 2 | | International Network of Kangaroo mother Care (INK) (Fundación Canguru) | https://fundacionc | - | 323 | | | APC-Colombia | 323 |

**Supplemental file 2 DATABASE OF WEBSITES AND REPOSITORIES**

For further exploration

| | Date | Name of website | URL | Search terms | # hits | Saved | File name | Comments | Identified for secondary screening |
|---|---|---|---|---|---|---|---|---|---|
| | 2021/07/20 | Kangaroo Mother Program | https://programac | - | 3 | Y | 2021-07-20 Colombia K | Website in addition to the INK one Found **3 documents for secondary screening - could not open one of them - Nathalie sent link for 3rd one** | 3 |
| | 2021/08/10 | fundacioncanguro.co | | - | 14 | | | Seems as if there has been a change in the website and previous documents are not accessible anymore **14 documents for secondary screening** | 14 |
| 3a | 2021/07/14 | Krisanne Larimer | http://www.geocities.com/roopage/kcresearch.html | | | **NOT FOUND** | | KC BIB: 3. Krissanne Larimer has a website for KC and the KC bib is available off this web site. The site is http://www.geocities.com/roopage and a list of Dr. Ludington's outcomes chart is at http://www.geocities.com/roopage/kcresearch.html. | |
| 3b | 2021/07/14 | Krisanne Larimer | http://www.prematurity.org/baby/kangaroo. | | **NO RELEVANT RESOURCES** | | | KC BIB: 3. Krissane Larimer also has another web site, and the document on it is Kangaroo Care Benefits. http://www.prematurity.org/baby/kangaroo.html | |
| 4 | 2021/07/14 | New York City health site | www.pathfinder.com/NY1/living/health/kan | | **COULD NOT ACCESS** | | | KC BIB: 4. www.pathfinder.com/NY1/living/health/kangaroo_baby_care This is New York city health site that reports where one can get Kangaroo Care in New York City and its outcomes. A very brief site. | |
| 5 | 2021/07/14 | Unnamed (Nils Bergman?) | | | **COULD NOT ACCESS** | | | KC BIB: 5. has some articles that you can request and the articles have been written by Nils Bergman. | |
| 6 | 2021/07/14 | See no. 2 | | | **NOT CONSULTED** | | | KC BIB: is a major KC Network email that is maintained by the Bogota group. It has many updates and should be checked regularly. It published as version of Dr. Ludington's KC bib. This site is maintained by Natalie Charpak and Natalie Charpak's email is | |
| 7 | 2021/07/14 | Closed down? | | | **NOT CONSULTED** | | | KC BIB: has Dr. Ludington's and Dr. Andersons' bibs on it. A Jan.29, 2010 note from Pat somewhere in the world says that this website is being used for dating now and it should be shut down. The new KMC group can be found in #7. | |
| 8 | 2021/07/14 | ??? | http://health.groups.yahoo.com/group/Kan | | **COULD NOT ACCESS** | | | KC BIB: 8. http://health.groups.yahoo.com/group/Kangaroo-Mother-Care/ is the website that has replaced #6 above as of Jan. 2010 | |

**Supplemental file 2 DATABASE OF WEBSITES AND REPOSITORIES**

For further exploration

| | Date | Name of website | URL | Search terms | # hits | Saved | File name | Comments | Identified for secondary screening |
|---|---|---|---|---|---|---|---|---|---|
| 9 | 2021/07/14 | Susan Ludington | http://fpb.case.edu | kangaroo care | 176 | Y | 2021-07-18 Case Weste | KC BIB: 9. http://fpb.case.edu/KangarooCare/biblio.shtm is the website that contains updated annotated bibliography of all publications about KC in the world that are known by Susan Ludington.<br>Could only access the first 91 hits - none relevant<br>Came across the Kangaroo Care Lab (https://case.edu/nursing/postgrad/research/research-studies-labs/kangaroo-care) - description of the US KC Institue - nothing relevant<br>There is also a Case Western University Kangaroo Care Lab - https://case.edu/search-results/?q=kangaroo+care&cx=004305171799132815236%253Aciq4c8b3yv4&ie=UTF-8 | |
| 10 | 2021/07/14 | Preemienews | http://preemienews.com | | COULD NOT ACCESS | | | KC BIB: 10. http://preemienews.com is a website that in July 2000 had an article on KC that reports the opinion of several doctors and developmental specialists on KC and all opinions are positive. | |
| 11 | 2021/07/14 | Lancet | http://news.bbc.co.uk/hi/english/newsid_18 | | COULD NOT ACCESS | | | KC BIB: 11. 1998 BBC. "Kangaroo Care Counters the Cold." This is a summary of Christensson's 1998 article in the LANCET. http://news.bbc.co.uk/hi/english/newsid_184000/184480.stm | |
| 12 | 2021/07/14 | Nils Bergman report | http://kangaroo.javeriana.edu.co/abstract4 | | GOT A SECURItY ALERT - COULD NOT AC | | | KC BIB: 12. 2000 Bergman, Nils. "Charge for the future of KC: A public health imperative." Available at http://kangaroo.javeriana.edu.co/abstract42.htm. This is a report of his presentation at the First International Kangaroo Care Conference held Oct. 23-25, 1998 in Baltimore, MD. | |
| 13a | 2021/07/14 | Nils Bergman webisite | www.kangaroomothercare.com | | NO RELEVANT RESOURCES | | | KC BIB: 13. 2003 Bergman, Nils. Kangaroo Mother Care website, listing his tour dates, the KMC Shop with videos, postcards, Kangacarrier Shirts for sale, and reference list from USIKC. Go to www.kangaroomothercare.com. He also has the website www.skintoskincontact.com | |
| 13b | 2021/07/14 | Nils Bergman webisite | www.skintoskincontact.com | | COULD NOT ACCESS | | | KC BIB: 13. 2003 Bergman, Nils. Kangaroo Mother Care website, listing his tour dates, the KMC Shop with videos, postcards, Kangacarrier Shirts for sale, and reference list from USIKC. Go to www.kangaroomothercare.com. He also has the website www.skintoskincontact.com | |
| 14 | 2021/07/14 | March of Dimes | www.marchofdime | kangaroo mother care<br>kangaroo | NO RELEVANT RESOURCES | | | KC BIB: 14. March of Dimes in 2005 started a prematurity campaign and developed a website that has much about the good of Kangaroo Care in it. Go to: www.marchofdimes.com/prematurity to see what they have. | |
| 15 | 2021/07/14 | NICU web page | http://vuneo.org-KangarooCare | | COULD NOT ACCESS | | | KC BIB: 15. NICU web page http://vuneo.org-KangarooCare | |

Supplemental file 2 DATABASE OF WEBSITES AND REPOSITORIES

For further exploration

| | Date | Name of website | URL | Search terms | # hits | Saved | File name | Comments | Identified for secondary screening |
|---|---|---|---|---|---|---|---|---|---|
| 16 | 2021/07/14 | India | http://www.kmcindia.com | | COULD NOT ACCESS | | | Landed on a website HugeDomains.com that says Kmcindia.com is for sale for $13,195<br>KC BIB: 16. India has a big initiative on Kangaroo Care and has its own website, http://www.kmcindia.com and it has content, overview of KC effects, and the goals to educate everyone in the country about KMC to improve its usage. Added 8/24/2007 | |
| 17 | 2021/07/14 | @MothersUtopia@Laura_K | www.obnurse35yrs.wordpress.com/tag/ski | | SITE NOT EXISTInG ANYMORE | | | Message on website: obnurse35yrs.wordpress.com is no longer available. The authors have deleted this site.<br>KC BIB: 17. @MothersUtopia@Laura_Keegan. OR through www.obnurse35yrs.wordpress.com/tag/skin-to-skin/<br>This is a website that is promotiong Kangaroo Care right after birith with every cesarean section. Mothers tell their stories here and it recommends that they go to see the HEALTH-INFO video which has a trailer that was posted on facebook. Accessed 9/11/2011. | |
| 18 | | Breastcrawl | http://www.breastcrawl.org | | NOT CONSULTED | | | KC BIB: 18.http://www.breastcrawl.org is the website to see a video of a fullterm newborn bieng placed between breasts and choosing which nipple to move towards and latch onto. The vido is from India and the infant remains wet the whole time several health professionals stand around and no cover on the baby's back, no drying off, no head cap, not a good example of thermoregulation, but the baby does move toward the nipple and latch on. Added 8/14/2007 | |
| 19 | | ??? | http://www.babygooroo.com/index.php/20 | | NOT CONSULTED | | | KC BIB: 19. http://www.babygooroo.com/index.php/2010/10/06/the-case-for-kangaroo-daddy-care. | |
| 20 | | Johnson & Johnson | http://www/jjpi.com | | NOT CONSULTED | | | KC BIB: 20. http://www/jjpi.com. This is the Johnson & Johnson Pediatric Institute website and in 2002, Johnson & Johnson produced a book for pediatricians and health professionals to disseminate for free that was called, "Skin-to-Skin: The Mother-Baby Package." | |
| 21 | 2021/07/14 | Nurtured by Design (Yamile Jackson) | https://thezaky.com/ | | COMMERCIAL SITE - NO RELEVANT RESO | | | KC BIB: 21. Kangaroo Care LINKEDIN Professional Group for KC discussions. Contact the moderator Yamile Jackson. Yamile is a Ph.D. Industrial Engineer who is a Certified Kangaroo Caregiver (2010). | |
| 22 | 2021/07/14 | The Miracle of Kangaroo Mother Care | www.themiracleofkangaroomothercare.coi | | COULD NOT ACCESS | | | KC BIB: 22. www.themiracleofkangaroomothercare.com . This website relates information on accessing the Roos T, and Roos N online book entitled The Miracle of Kangaroo Mother Care. Rare Inspirational Stories of Infant Survival for Every Parent and Every Baby. See Roos citation in first section. This website also gives access to the "Kangaroo Mohter Care: The Benefits for Your Full Term and Premature Baby." 2-page pamphlet/information sheet that is listed in the main section of the bibliography under Lawm, J. (2011) and also under the Pamphlet section as Lawn, J. (2011). | |

Supplemental file 2 DATABASE OF WEBSITES AND REPOSITORIES

For further exploration

| | Date | Name of website | URL | Search terms | # hits | Saved | File name | Comments | Identified for secondary screening |
|---|---|---|---|---|---|---|---|---|---|
| 23 | | DrBarbCNM.com | No URL given | | NOT CONSULTED | | | KC BIB: 23. DrBarbCNM.com This is a website that advertises the three CDs of Dr. Barbara Morrison giving talks about Kangaroo Care with full term infants that can be purchased. Her 2007 talk, Kangaroo Care: Nature's Best for your Baby, which is 20 minutes long, is available for $25.00. | |
| 24 | | www.healthed.cc | | | NOT CONSULTED | | | KC BIB: 24. www.healthed.cc This is the website for getting pamphlets about KC with full term infants (one is called How To Hold Your Bay Skin-to-Skin and the other is called The First Hour After Birth: A Baby's 9 Instinctive Stages. | |
| 25 | | International Network of Kangaroo Mother Care (INK) | No URL given - see 29 below | | DUPLICATE | | | KC BIB: 25. International Network of Kangaroo Mother Care (INK) has a website that started in summer 2013. Each month the blog changes reporting what is happening in countries with KMC being practiced. | |
| 26 | 2021/07/14 | Improving Birth Coalition | http://www.motherfriendly.org | | NO RELEVANT RESOURCES | | | KC BIB: 26. http://www.motherfriently.org | |
| 27 | 2021/07/14 | WHO | http://who.int/reproductive-health/publicatio | | COULD NOT ACCESS | | | KC BIB: 27. http://who.int/reproductive-health/publications/KMC/text.pdf | |
| 28 | 2021/07/14 | Prematurity | http://Prematurity.org | | NO RELEVANT RESOURCES | | | Mainly resources for parenting<br>KC BIB: 28. http://Prematurity.org | |
| 29 | 2021/07/14 | International Network of Kangaroo Mother Care (INK) | www.INKmc.net | | COULD NOT ACCESS | | | Message on website: It is possible you have reached this page because: (a) The IP address has changed; (b) There has been a server misconfiguration; (c) The site may have moved to a different server<br>KC BIB: 29. The International Network of Kangaroo Mother Care (INK for International Network of Kagarooing) has its very own website as of Oct. 4, 2013. Go to www.INKmc.net for the home page. | |
| 30 | 2021/07/14 | YOUR RESOURCE FOR CO | www.support4nicuparents.org | | COULD NOT ACCESS | | | Family-centered developmental Care - nothing relevant<br>KC BIB: 30. www.support4nicuparents.org has many articles from National Perinatal Association guidelines that speak to Kangaroo Care extensively. | |
| 31 | | United State Institute for Kangaroo Care - see number 1 | | | DUPLICATE | | | KC BIB: 31. United State Institute for Kangaroo Care's website is www.kangaroocareusa.org and it features videos to watch, lectures that can be ordered to be given, position papers, and the flull Kangaroo Care Bibliography.I79 | |

**Supplemental file 2 DATABASE OF WEBSITES AND REPOSITORIES**

For further exploration

| | Date | Name of website | URL | Search terms | # hits | Saved | File name | Comments | Identified for secondary screening |
|---|---|---|---|---|---|---|---|---|---|
| 32 | | Healthy Children Project Inc. (Skin to skin Practical advice to staff) | www.skin2skin.org | | NO RELEVANT RESOURCES | | | KC BIB: 32. www.skin2skin.org. This is the website of Children Project, Inc. 327 Quaker Meeting House Road, East Sandwich, MA 02537 phone: 508-888-8044; fax: 508-888-8050, This website has 6 headings and the HOME page shows pictures and tells reader why SSC is important ad vital, listing benefits to baby and to mother (limited list of benefits, however), under 9 STAGES page, it has pictures of the babies in each of Widstrom's nine stages of getting to the breast. These nine pictures are available for sell under the RESOURCES page. The RESEARCH page relates the latest research about KMC in Gulu, Uganda and has several videos to watch. This section also tells you about the two publications of studies by Brimdyr "The Association between common labor drugs and sucking when skin to skin during the first hour after birth" in Birth: Issues in Perinatal Care, 2015 (on the bib above), and Kajsa Brimdyr's (Ph.D. Lctation consulatant) and Karen Cadwell's "An Implementation Algorithm to Improve skin to skin practice in the first hour after birth" (also on bib aboave). The research team of Healthy Children Project is comprised of K Brimdyr, Ann-Marie Widstrom, Kristin Svensson RN, Midwife, MD, and Karin Cadwell, RN, PhD.., IBCLC, FAAN. Ther RESOURCE page inclues videos of the the film skin to skin in the first hour after birth, the Magical Hour (same content, but for parents), and Happy Birthday (which shows skin to skin contact), and concludes with sales advertisements forTEAR OFF PADS ENTITLED " HOW TO HOLD YOUR BABY SKIN-TO-SKIN," " THE FIRST HOUR AFTER BIRTH," AND "A BABY'S 9 INSTINCTUAL STAGES." And how you can order them. The REFERENCE page includes some of the articles on the KC bib and a few government websites, The CONTACT pages gives the address, phone, fax and email address of The Healthy Children Project in Massachusetts. | |
| 33 | | British Association of Perinatal Medicine | www.bapm.org | | 3 | N | - | **3 documents on shared decision making, service quality indicators and serivce standards for secondary screening** KC BIB: 33. www.bapm.org is a british perinatal association that published a 36 page document entitled "Guidelines for the investigation of Newborn Infants who Suffer from Sudden Unexpected Postnatal Collapse in the First Week of Life. Recommendations from a Professional Group on Sudden Unexpected Postnatal Collapse, March 2011" under Beecher on the bib above. | 3 |
| | 2021/07/15 | USAID Health Research Program | https://www.harpr | kangaroo mother care | 0 | Y | 2021-07-15 USAID Heal | Stumbled upon website during a webinar on collaborative learning | |
| | 2021/07/19 | Evidence Action | https://www.evide | kangaroo mother care | 0 | N | - | Had an interview once with them | |
| | 2021/07/20 | KMC Foundation India | https://kmcfounda | - | 11 | Y | 2021-07-20 Webpage K | **11 newsletters downloaded for secondary screening** | 11 |

**Supplemental file 2 DATABASE OF WEBSITES AND REPOSITORIES**

For further exploration

| Date | Name of website | URL | Search terms | # hits | Saved | File name | Comments | Identified for secondary screening |
|---|---|---|---|---|---|---|---|---|
| 2021/07/23 | USAID Maternal and Child Survival Program (MCSP) | https://www.mcsp | kangaoo more care | 32 | N | - | **Downloaded 36 documents for secondary screening** | 36 |
| 2021/07/23 | Jhpiego | https://resources.j | kangaroo mother care | 28 | Y | 2021-07-23 Jpiego | Many resources taken over from elsehwere, also published articles<br>**4 MCHIP documents downloaded for secondary screening** | 4 |
| 2021/07/24 | USAID (maternal and child health) | https://www.usaid | kangaroo mother care | 0 | Y | 2021-07-24 U.S. Agency | Also hand-searched the titles under Global Health - Maternal and Newborn Health<br>Mostly documents sponsored by USAID that are published by another agency (e.g. WHO, UNICEF)<br>Reference to their flagship programmes are followed up in the separate URLs provided | |
| 2021/07/24 | USAID Maternal and Child Health Integrated Program (MCHIP) (Prematurity and LBW) | https://www.m | kangaroo mother care | 9 | Y | 2021-07-24 MCHIP Technical Resources<br>2021-07-27 MCHIP Country Programs | Some of the resources already identified through other searches (e.g. Jpiego)<br>**15 additional documents downloaded for secondary screening**<br>**1 document did not want to download - marked with an X in folder for follow-up with Goldy**<br>**11 end of project country reports that mention KMC (25 country reports with nothing on KMC)** | 26 |
| 2021/07/24 | USAID Maternal and Child Health Integrated Program (MCHIP) (Newborn health) | https://www.mchip | - | 0 | N | - | Only duplicates from the search under Prematurity and LBW | |
| 2021/07/24 | USAID Every Preemie--Scale | https://www.every | - | 2 | N | - | Used personal knowledge of the projet to access documents<br>**2 documents for secondary screening** | 2 |
| 2021/07/24 | Align MNH | https://www.alignr | - | - | N | - | | |
| 2021/07/24 | NEST360° (Newborn Essential Solutions and Technologies) | | - | - | Y | 2021-07-24 NEST360 to | NEST toolkit will be available in November 2021<br>Health systems strengthening organsed by the health systems building blocks (plus infection prevention and control as a separate block) | |
| 2021/07/24 | Clinton Health Access Initiative (CHAI) | https://www.clinto | kangaroo | 1 | Y | CHAI KMC is cue from r | Blog - story of a women - **1 for secondary screening** | 1 |
| 2021/07/24 | Jhpiego Access to Health (ACCESS) | https://www.jhpieg | - | - | Y | 2021-07-24 Access to H | No resources isted - only a short summary of ACCESS | |
| 2021/07/24 | JSI Research & Training Institute, Inc. | https://www.jsi.co | kangaroo mother care | 1025 | Y | 2021-07-24 John Snow | Appears as a small window that cannot be opened in full screen<br>Went through the first 60 hits - after 20 the hits were no longer "kangaroo mother care" as one term<br>**15 documents downloaded for secondary sceening** | 15 |
| 2021/07/24 | UKAID | https://www.ukaid | kangaroo | 0 | N | - | | |
| 2021/07/24 | Bill & Melinda gates Foundation | https://www.gates | kangaroo | 2 | Y | 2021-07-24 Bill & Melind | 2 speeches in which K(M)C is mentioned in passing<br>12 commited grants listed that has KMC somewhere in the documents | |

**Supplemental file 2 DATABASE OF WEBSITES AND REPOSITORIES**

(yellow) For further exploration

| Date | Name of website | URL | Search terms | # hits | Saved | File name | Comments | Identified for secondary screening |
|---|---|---|---|---|---|---|---|---|
| 2021/07/24 | USAID MOMENTUM (Moving Integrated, Quality Maternal, Newborn, and Child Health and Family Planning and Reproductive Health Services to Scale) | https://www.usaid | kangaroo mother care | 0 | N | - | Part of USAID website - same documents listed | |
| 2021/07/24 | Healthy Newborn Network (HNN) | https://www.health | **Filter: kangaroo mother care** | 173 | Y | 2021-07-24 Healthy New | Also most of the open access adrticles related to KMC<br>**97 documents downloaded for secondary screening**<br>**3 documents could not download - try again later** | 97 |
| 2021/07/25 | Healthy Newborn Network (HNN) | https://www.health | **Filter: scale-up** | 231 | Y | 2021-07-25 Healthy New | Revisit later when writing about scale-up<br>**35 documents downloaded that contain possible themes - do not inlcude in initial analysis** | |
| 2021/07/26 | KMC Acceleration Partnership Community of Practice (KAP COP) | https://knowledge | - | 8 | Y | 2021-07-26 KAP COP K | **Discussion consolidated in 1 Word document - condense for secondary analysis**<br>**8 documents downloaded for secondary analysis** | 9 |
| 2021/07/26 | PATH | https://www.path.o | kangaroo | 25 | Y | 2021-07-26 PATH | Could not access documents dated before 2017<br>**Humn Milk Bank Toolkit (counted as 1 document)**<br>**16 more documents downloaded for secondary screening** | 17 |
| 2021/07/27 | Nutrition International | https://www.nutriti | kangaroo | 24 | Y | 2021-07-27 Nutrition Inte | Mostly field reports and blogs about individual cases/women<br>**All 24 documents downloaded for secondary screening** | 24 |
| 2021/07/27 | Partners in Health | https://www.pih.or | kangaroo | 18 | Y | 2021-07-27 Partners In | Could not access all hits - for some there was a donation form which I could not get past<br>**12 documents downloaded for secondary screening** | 12 |
| 2021/07/27 | UNICEF | https://www.unice | kangaroo mother care | 10 | Y | 2021-07-27 UNICEF | **11 documents downloaded for secondary screening** | 11 |
| 2021/07/27 | Médecins sans Frontières (MSF) | https://www.msf.o | kangaroo mother care | 8 | Y | 2021-07-27 MSF | **6 documents downloaded for secondary screening** | 6 |
| 2021/07/27 | Save the Children (Resource centre) | https://resourcece | kangaroo mother care (free text) | 150 | Y | 2021-07-27 Save the Ch | Quite a numer of published articles - also documents already downloaded from other sites - there may be a few duplicates in this search - 13 documents with KMC or preterm/LBW in title<br>**35 documents downloaded for secondary screening** | 35 |
| 2021/08/10 | National Institute for Health Care and Excellence (NICE) | https://www.evide | kangaroo guidelines | 87 | Y | 2021-08-10 NICE | Most documents are published papers and UN documents<br>**Downloaded 2 documents for secondary screening** | 2 |
| 2021/08/14 | First Embrace | | - | ??? | Y | 2021-08-14 The first Em | Difficult to access the resources<br>**Added 4 documents for secondary screening** | 4 |

**Supplemental file 2 DATABASE OF WEBSITES AND REPOSITORIES**

For further exploration

| Date | Name of website | URL | Search terms | # hits | Saved | File name | Comments | Identified for secondary screening |
|---|---|---|---|---|---|---|---|---|
| 101 websites & repositories consulted | | | | | | | | 701 |
| 701 documents | | | | | | | | |
| 42 | Documents received from individuals | | | | | | | 42 |
| 10 | Own knowledge | | | | | | | 10 |

**TOTAL NUMBER OF DOCUMENTS IDENTIFIED FOR SECONDARY SCREENING** 753

**HITS (at least):** 52787

(Two sites unknown number of hits)
