## Supplemental file 3 for "Programmatic implementation of kangaroo mother care: a systematic synthesis of grey literature"

### Categories included in the MS Excel database, with screenshots

1. ID
2. Title (Open-ended)
3. Publishing 'authority' (Drop down: Independent author/s; Media house; Funding agency; Development agency; Government [specify]; Other [specify])
   (a) If GOVERNMENT, specify
   (b) If OTHER, specify
4. Type of document: (Drop down: Book; Report; Guidelines; Conference abstract; Conference presentation; Conference proceedings; Scientific article (peer-reviewed); Scholarly article (non-peer-reviewed or uncertain) (e.g. Perspective); 'Official' media report; Blog; Other [specify])
   (a) If OTHER, specify
5. Authors Compilers (if listed)
6. Donor / Development support (Dropdown: YES / NO / UNSURE)
   If YES: (a) Linked to a time-limited or once-off project? YES / NO / UNSURE
   (b) Specify further (open-ended - will be clustered at a later stage if needed)
7. Country/ies (open-ended – will be clustered at a later stage if needed) If no country, use "Unknown" or "General" or another term that may emerge as relevant during the review
8. KMC coverage mentioned? YES / NO
   If YES: (a) Page number(s)
   (b) Key issues mentioned (if applicable)
   (c) Grading of document regarding coverage:
   A = quantitative information on HIGH KMC coverage (> 50%)
   B = quantitative information on ANY KMC coverage
   C = mention of HIGH coverage without quantitative information
   D = mention of ANY coverage without quantitative information
   E = NO mention of coverage
9. KMC implementation mentioned? YES / NO
   If YES: (a) Page number(s)
   (b) Key issues mentioned
   (c) Health system interventions mentioned? YES / NO/ UNSURE
   (i) If YES, more detailed analysis needed? YES / NO/ UNSURE
   - If YES or UNSURE, mark the ID cell in RED
10. KMC improvement/strengthening mentioned? YES / NO
    If YES: (a) Page number(s) and key issues mentioned
    (b) Key issues mentioned
    (c) Health system interventions mentioned? YES / NO / UNSURE
    (i) If YES, more detailed analysis needed? YES / NO / UNSURE
    - If YES or UNSURE, mark the ID cell in RED
11. KMC scale-up mentioned? YES / NO
    If YES: (a) Page number(s) and key issues mentioned
    (b) Key issues mentioned
    (c) Health system interventions mentioned? YES / NO / UNSURE
    (i) If YES, more detailed analysis needed? YES / NO / UNSURE
    - If YES or UNSURE, mark the ID cell in RED
    (d) Grading of document regarding health interventions related to KMC implementation and scale-up:
    A = detailed information on MORE THAN ONE COMPONENT of health interventions
    B = detailed information on ONLY ONE COMPONENT of health interventions
    C = mention MORE THAN ONE COMPONENT of health interventions

D = mention ONLY ONE COMPONENT of health interventions
E = NO mention of any components of health interventions

12. Quality: KMC initiation mentioned? YES / NO / UNSURE
    If YES or UNSURE: (a) Describe:
13. Quality: KMC dose (e.g. hours per day) mentioned? YES / NO / UNSURE
    If YES or UNSURE: (a) Describe:
14. Quality: KMC duration (number of days) mentioned? YES / NO / UNSURE
    If YES or UNSURE: (a) Describe:
15. Availability of KMC services (number of facilities providing KMC) mentioned? YES / NO / UNSURE
    If YES: (a) Numbers mentioned? YES / NO
    (i) If YES, describe (including if any comparison with total number of facilities with maternity and neonatal services):
16. First analysis completed? YES (when done) (Mark cell in GREEN) (Continue with an in-depth analysis of documents marked red in the ID cell)

### EXAMPLES OF SCREENSHOTS

#### (AB) Abstracts (also posters, summaries & working groups)

| | A | B | C | F | G | H | I | M | N | O | P | Q | R |
|---|---|---|---|---|---|---|---|---|---|---|---|---|---|
| 1 | **1 ID** | **2 Title** | **3 Name of conference** | **Date published/presented** | **4 Document type** | **4a Other (specify)** | **5 Authors/ compilers** | **7 Country/ies** | **8 Coverage mentioned** | **8a Page #** | **8b Key issues** | **8c Grading** | **9 Implementation mentioned** |
| 33 | AB31a | Workshop on Kangaroo Mother Care: lessons | KMC Conference, Trie | 2016 | Report | Working group | Pr Djamil LEBA | Algeria | YES | 1 | Low facility-based cove | D | YES |
| 34 | AB31b | ROUND TABLE: BARRIERS AND ENABLERS OF CO | KMC Conference, Trie | 2017 | Report | Round table | Pr Djamil LEBA | Algeria | NO | - | - | E | YES |
| 35 | AB32 | Brazil | KMC Conference, Trie | 2016 | Report | Working group | No name | Brazil | YES | 1-2 | Services & beds | C | YES |
| 36 | AB33a | CONSTRAINTS AND BARRIERS TO IMPLEMENTA | KMC Conference, Trie | 2016 | Report | Working group | NOUTCHOGOU | Cameroon | NO | - | - | E | YES |
| 37 | AB33b | Diagrams Laquintinie Hospital | KMC Conference, Trie | 2017 | Report | Working group | No name | Cameroon | NO | - | - | E | NO |
| 38 | AB34 | COLOMBIA | KMC Conference, Trie | 2016 | Report | Working group | No name | Colombia | YES | 1-2 | Population-based: 80% | A | YES |
| 39 | AB35 | Enablers and Challenges of Kangaroo Mother | KMC Conference, Trie | 2016 | Report | Working group | Shashi N. Vani | India | YES | 3 | Facility-based: 13.2% | B | YES |
| 40 | AB36a | KMC state of the art - Italy | KMC Conference, Trie | 2016 | Report | Working group | No name | Italy | NO | - | - | E | YES |
| 41 | AB36b | Rountable less successful countries, 14 Nove | KMC Conference, Trie | 2016 | Report | Round table | Immacolata Da | Italy | NO | - | - | E | YES |
| 42 | AB37 | KMC Workshop Italy 2016 | KMC Conference, Trie | 2016 | Report | Working group | (Mantoa Mock | South Africa | NO | - | - | E | YES |
| 43 | AB38 | Group work on enablers and barriers, 14 Nov | KMC Conference, Trie | 2016 | Report | Working group | (Mantoa Mock | SSA (Cameroon, | NO | - | - | E | YES |
| 44 | AB39 | Application of KMC in Sweden | KMC Conference, Trie | 2016 | Report | Working group | Kerstin Hedber | Sweden | INDIRECTLY | 1-2 | Services | E | YES |
| 45 | AB40 | Kangaroo mother care in Viet Nam: an overvie | KMC Conference, Trie | 2016 | Report | Working group | MCH Dept., Mi | Vietnam | YES | 2 | Facility-based coverage | B | YES |
| 46 | AB41 | Group work on enablers and barriers, 14 Nov | KMC Conference, Trie | 2016 | Report | Working group | No name | Brazil, India | NO | - | - | E | YES |
| 47 | AB42 | Group work on enablers and barriers, 14 Nov | KMC Conference, Trie | 2016 | Report | Working group | No name | Italy, Sweden, F | NO | - | - | E | YES |
| 48 | AB43 | Work group on enablers and barriers, 14 Nov | KMC Conference, Trie | 2016 | Report | Working group | No name | Pakistan, Mada | NO | - | - | E | YES |
| 49 | AB44 | Group workshop on enablers and barriers, 14 | KMC Conference, Trie | 2016 | Report | Working group | Nathaniel Foot | Vietnam, Colom | NO | - | - | E | YES |
| 50 | AB45 | Training workshop | KMC Conference, Trie | 2016 | Report | Working group | N Charpak, Ma | Colombia / USA | NO | - | - | E | NO |
| 51 | | | | | | | | | | | | | |
| 52 | 16 | | | | | | | | | | | | |
| 53 | 32.7% | | | Book | 0 | | **REPORT** | | 7 | YES | A | 1 | 41 |
| 54 | | | 36.73% | Report | 18 | 2 | Round table | | 34 | NO | B | 4 | 7 |
| 55 | | | 46.94% | Conf abstract | 23 | 16 | Working groups | | 0 | UNSURE | C | 1 | |
| 56 | | | | Conf presentati | 0 | | | | 0 | N/A | D | 6 | |
| 57 | | | | Peer-reviewed a | 0 | | | | 7 | INDIRECTLY | E | 36 | |
| 58 | | | | Article non-peer | 0 | | | | 1 | Missing | Missing | 1 | 1 |
| 59 | | | | Media report (o | 0 | | | | **49** | **Total** | **Total** | **49** | **49** |
| 60 | | | | Blog | 0 | | | | | | | | |
| 61 | | | 16.33% | Other | 8 | | **OTHER** | | | | | | |
| 62 | | | | **Total** | **49** | 1 | Conference summary | | | | | | |
| 63 | | | | | | 7 | Poster | | | | | | |
| 64 | | | | | | | | | | | | | |
| 65 | | | | | | | | | | | | | |

(AB) Abstracts (49) | (AS) Assessments (31) | (R) Reports (53) | (SP) Slides & pres (60) | (SC) Scal ...

#### (AS) Assessments, evaluations and reviews

| | A | B | N | O | P | Q | R | S | T | U | V | W | X |
|---|---|---|---|---|---|---|---|---|---|---|---|---|---|
| 1 | 1 ID | 2 Title | 8 Coverage mentioned | 8a Page # | 8b Key issues | 8c Grading | 9 Implementation mentioned | 9a Page # | 9b Key issues | 9c H.syst interventions mentioned | 9ci Analysis needed | 10 Improve/Strengthen (mentioned) | 10a Page # |
| 2 | AS1 | REVIEW OF KANGAROO MOTHER CARE IMPLEN | YES | 8 | Services | D | YES | Various pa | Stages of change mod | YES | YES | YES | Throughout |
| 3 | AS2 | To explore the feasibility of introduction and | NO | - | - | E | YES | 11, 21-32 | Service Provision Ass | INDIRECTLY | UNSURE | NO | - |
| 4 | AS3 | BANGLADESH National Newborn Health Situat | NO | - | - | E | YES | 48 | Summary table of im | YES | YES | INDIRECTLY | - |
| 5 | AS4 | Executive summary Review of Kangaroo Moth | NO | - | - | E | YES | 1-2 | Recommendations fo | YES | YES | YES | 1-2 |
| 6 | AS5 | Kangaroo Mother Care | NO | - | - | E | YES | | Conclusion: "KMC wo | NO | NO | NO | - |
| 7 | AS6 | An Assessment of the Status of Kangaroo Mot | YES | 11, 17, 20, | Geographic coverage, c | D | YES | Throughout | Analysis according to | YES | YES | YES | Throughout |
| 8 | AS7 | The feasibility and acceptability of Kangaroo | YES | 8 | Definition for populatic | D | YES | Throughout | Perceptions of famili | YES | YES | INDIRECTLY | 24-5 |
| 9 | AS8 | A Health System Bottleneck Analysis of Care a | YES | 14, 20 | Staff coverage, coverag | D | NO | | KMC not discussed ir | INDIRECTLY | YES | INDIRECTLY | |
| 10 | AS9 | Situation analysis of newborn health in Tanza | YES | 19 | Coverage & quality of s | D | YES | Various pa | Nothing on KMC in da | INDIRECTLY | UNSURE | INDIRECTLY | |
| 11 | AS10 | FIELD ACTION REPORT Challenges and Lesson | YES | Multiple pa | Coverage & quality, cov | C | YES | 16 | Country progress wit | YES | YES | YES | 17 |
| 12 | AS11 | Survey of kangaroo mother care in North Wes | YES | 1,6 | Coverage of services | C | YES | Multiple sl | Stages of change mod | YES | YES | YES | Multiple sl |
| 13 | AS12 | Trip report and action points CHAD | YES | 3 | Universal health covera | D | YES | Various pa | Stages of change mod | YES | YES | YES | 10-15 |
| 14 | AS13 | Trip report and action points TOGO | YES | 14 | Population-based | D | YES | Various pa | Stages of change mod | YES | YES | YES | 14-18 |
| 15 | AS14 | Implementation and evaluation of kangaroo r | YES | Multiple pa | Population-based | D | YES | Various pa | Check in further anal | YES | YES | YES | Throughout |
| 16 | AS15 | Retrospective Evaluation of Kangaroo Mother | YES | 7,20 | Vague | D | YES | Various pa | Check in further anal | YES | YES | YES | Various pa |
| 17 | AS16 | Tracking implementation progress of Kangarc | NO | - | - | E | YES | Various pa | Check in further anal | YES | YES | YES | 7-8 |
| 18 | AS17 | EVALUATION OF KANGAROO MOTHER CARE SEI | NO | - | - | E | YES | Various pa | Check in further anal | YES | YES | YES | 34-36 |
| 19 | AS18 | EVALUATION OF KANGAROO MOTHER CARE SEI | NO | - | - | E | YES | Various pa | Check in further anal | YES | YES | YES | 25-27 |
| 20 | AS19 | EVALUATION OF KANGAROO MOTHER CARE SEI | YES | 18 | Services | D | YES | Various pa | Check in further anal | YES | YES | YES | 27-30 |
| 21 | AS20 | EVALUATION OF KANGAROO MOTHER CARE SEI | YES | 12 | Coverage of newborn ca | D | YES | Various pa | Check in further anal | YES | YES | YES | 32-33 |
| 22 | AS21 | The implementation and scale up of facility-b | YES | 1,5,27,28,3 | Coverage of services; AI | D | YES | Various pa | Check in further anal | YES | YES | YES | 48-50 |
| 23 | AS22 | EVALUATION OF KANGAROO MOTHER CARE SEI | YES | 42 | Services | D | YES | Various pa | Check in further anal | YES | YES | YES | 38-41 |
| 24 | AS23 | Kangaroo Mother Care in the Philippines: Ass | YES | 30 | Health insurance cover | D | YES | Various pa | Check in further anal | YES | YES | YES | 31-32 |
| 25 | AS24 | Review of Kangaroo Mother Care in Indonesia | YES | 43 | Population-based (also | D | YES | Various pa | Check in further anal | YES | YES | YES | 61-64 |
| 26 | AS25 | Exploration of Opportunities for Facility-base | YES | 35 | MNCH services | D | YES | Various pa | Check in further anal | YES | YES | YES | 34-39 |
| 27 | AS26 | Situation Analysis Report on the Introduction | YES | 1,41 | KMC services | D | [illegible] | Various pa | Check in further anal | YES | YES | YES | 45-48 |
| 28 | AS27 | PUBLIC HOSPITAL-BASED CARE OF SMALL NEWI | YES | 7 | Services | E | YES | Throughout | Check in further anal | YES | YES | YES | 3-7 |
| 29 | AS28 | PUBLIC HOSPITAL-BASED CARE OF SMALL NEWI | YES | Multiple pa | Services | D | YES | Throughout | Check in further anal | YES | YES | YES | 101-110 |
| 30 | AS29 | Establishing a Centre of Excellence for Mater | NO | - | - | E | YES | Throughout | Research centre of ex | INDIRECTLY | UNSURE | INDIRECTLY | |
| 31 | AS30 | AN EVALUATION OF THE ESTABLISHMENT OF TH | NO | - | - | E | YES | Throughout | Research centre of ex | INDIRECTLY | UNSURE | INDIRECTLY | |
| 32 | AS31 | Rapid Health Facility Assessment on Service A | NO | - | - | E | YES | 2 | Reasons for not prov | YES | YES | YES | 3-4 |
| 33 | | | | | | | | | | | | | |
| 34 | 24 | Independent author/s | | | | | | | | | | | |
| 35 | 77.4% | Media House | 21 | YES | A | 0 | 30 | YES | YES | 25 | 26 | 23 | YES |

(AB) Abstracts (49) | (AS) Assessments (31) | (R) Reports (53) | (SP) Slides & pres (60) | (SC) Scal ...

#### (R) Reports, briefs, newsletters & blogs

| | A | B | W | X | Y | Z | AA | AB | AC | AD | AE | AF | AG |
|---|---|---|---|---|---|---|---|---|---|---|---|---|---|
| 1 | 1 ID | 2 Title | 10 Improve/Strengthen (mentioned) (KMC services) | 10a Page # | 10bKey issues | 10c H.syst interv. (mentioned) | 10ci Analysis needed | 11 Scale-up (mentioned) | 11a Page # | 11bKey issues | 11c H.syst interv. (mentioned) | 11ci Analysis needed | 11d Grading |
| 2 | R1 | Improving Newborn Survival (INS) project: A r | YES | 9 | Improvement of QoC o | NO | NO | NO | - | - | NO | NO | C |
| 3 | R2 | Kangaroo Mother Care: to prevent deaths due | NO | - | - | NO | NO | INDIRECTLY | 15 | See Remarks 50% by 202 | NO | NO | E |
| 4 | R3 | Experiences in Kangaroo Mother Care in Five | YES | 11 | Re-implementation de | YES | YES | YES | 3; 5; 8 | Scale-up plans for 3 cou | INDIRECTLY | YES | C |
| 5 | R4 | Iraq hospitals start "Kangaroo Mother Care" | NO | - | - | NO | NO | YES | 6 | Target another hospital ( | NO | NO | E |
| 6 | R5 | IMPROVING OUTCOMES OF PREMATURE AND L | YES | 2 | Re-implementation (tr | YES | YES | YES | 2 | "HCI provides technical a | INDIRECTLY | YES | C |
| 7 | R6 | Khanda ndi Mphatso Campaign Report | NO | - | Strengthening of colla | YES | UNSURE | NO | - | - | NO | NO | B |
| 8 | R7 | Development of a National Routine Reporting | YES | | Pilot improved KMC fo | YES | UNSURE | YES | 3 | Roll-put of register to res | YES | YES | B |
| 9 | R8 | Improving Uptake of Skin-to-Skin Practice for | YES | Various pa | Hypothesis that specia | YES | UNSURE | YES | 7 | Scale-up of wrapper | YES | YES | B |
| 10 | R9 | Promoting Kangaroo Mother Care in Selected | NO | - | - | NO | NO | NO | - | - | NO | NO | B |
| 11 | R10 | First Embrace: First biennial progress report | INDIRECTLY | - | Status diagrams for ea | NO | NO | INDIRECTLY | - | Benchmarks for EENC sca | NO | NO | A |
| 12 | R11 | Second biennial progress report (2017-2017) | INDIRECTLY | - | Status diagrams for ea | NO | NO | YES | 33-34 | Benchmarks for EENC sca | YES | YES | A |
| 13 | R12 | PROBLEMS AND SOLUTIONS FOR THE IMPLEME | YES | Whole doc | Solutiosn to problems | YES | YES | NO | - | - | NO | NO | C |
| 14 | R13 | JSI's contribution under Vriddhi for implemen | YES | 2 | Strengthening system | YES | YES | YES | | The ultimate aim of the t | YES | YES | C |
| 15 | R14 | VRIDDHI PROJECT Improving Maternal, Newbo | YES | 5-6 | Strengthening system | YES | YES | YES | 5-6 | Scale-up in 2 states | YES | YES | C |
| 16 | R15 | Kangaroo Mother Care Saves Newborns | YES | 3 | Way forward (nationa | YES | NO | NO | - | - | NO | NO | C |
| 17 | R16 | Kangaroo Mother Care: Opportunities for Nat | NO | - | - | NO | NO | YES | 4 | Recommendations for Fu | YES | YES | A |
| 18 | R17 | Kangroo Mother Care in Bangladesh | YES | 6-7 | Challenges, lessons ar | YES | YES | YES | Whole doc | Government commitmen | YES | YES | A |
| 19 | R18 | Kangroo Mother Care in Ethiopia | YES | 4-5 | Challenges, lessons ar | YES | YES | YES | Whole doc | Gaps for scale-up and re | YES | YES | A |
| 20 | R19 | Kangroo Mother Care in India | YES | 5 | Challenges, lessons ar | YES | YES | YES | 1 | "MOHFW piloted an inte | YES | YES | A |
| 21 | R20 | Kangroo Mother Care in Malawi | YES | 5 | Challenges, lessons ar | YES | YES | YES | 5 | Factors that facilitated s | YES | YES | C |
| 22 | R21 | Kangroo Mother Care in Nigeria | YES | 5-6 | Challenges, lessons ar | YES | YES | YES | 3,6 | Govt not funding scale-u | YES | NO | D |
| 23 | R22 | Kangroo Mother Care in Rwanda | YES | 3-4 | Challenges, lessons ar | YES | YES | YES | 1,4 | Factors that facilitated s | YES | YES | C |
| 24 | R23 | KMC Acceleration Partnership Community of | NO | - | - | NO | NO | YES | 15 | To be discussed in next r | NO | NO | C |
| 25 | R24 | Kangaroo Mother Care Acceleration Partnersl | YES | Various pa | Improve quality of ser | YES | YES | YES | Various pa | One of aims of meeting = | NO | NO | C |
| 26 | R25 | KMC Acceleration Partnership Community of | NO | - | - | NO | NO | YES | 16 | To be discussed in next r | NO | NO | C |
| 27 | R26 | Africa KAP Supplemental Report December 15 | YES | 9, 59, 103, | Sustainability; docum | YES | YES | YES | 54-59 | Summary of KMC acceler | YES | YES | C |
| 28 | R27 | Asia KAP Supplemental Report December 10- | YES | 212, 230, | Improved survival & l | YES | YES | YES | Multiple sl | Bangladesh (pp158-162) | YES | YES | C |
| 29 | R28 | Meeting Slides KMC Acceleration Partnership | YES | 114, 490, 5 | Family participative c | YES | YES | YES | Multiple sl | Reports on numbers of h | YES | YES | C |
| 30 | R29 | Neonatal Alliance Toolkit | UNSURE | - | - | NO | NO | YES | Various pa | Reference to interventior | YES | YES | C |
| 31 | R30 | Prematurity and Low Birth Weight | NO | - | - | NO | NO | YES | Whole doc | Only reference to MCHIP | NO | NO | E |
| 32 | R31 | Strengthening Small Baby Care at Thanlyin G | YES | - | Strengthening of small | NO | NO | NO | - | - | NO | NO | C |
| 33 | R32 | Brief 3: Facility Readiness and Initiation of K | UNSURE | - | - | NO | NO | NO | - | - | NO | NO | C |
| 34 | R33 | GLOBAL HEALTH NEWBORN LEGACY | NO | - | - | NO | NO | YES | 7 | - | NO | NO | E |

(AB) Abstracts (49) | (AS) Assessments (31) | (R) Reports (53) | (SP) Slides & pres (60) | (SC) Scal ...

#### (SP) Slides & conference presentations

| | A | B | AE | AF | AG | AH | AI | AJ | AK | AL | AM | AN | AO |
|---|---|---|---|---|---|---|---|---|---|---|---|---|---|
| 1 | 1 ID | 2 Title | 11c H.syst interv. (mentioned) | 11ci Analysis needed | 11d Grading | 13 Qual initiation | 13a Describe | 14 Qual dose | 14a Describe | 15 Qual duration | 15a Describe | REMARKS | 16 Anal compl EXCEL |
| 44 | SP43 | Accelerating scale up of KMC, what will it t | YES | YES | C | NO | | NO | | NO | | Ends with ENAP | |
| 45 | SP44 | Understanding Barriers to Practicing Kang | NO | NO | E | NO | | NO | | NO | | Qualitative study with parents in Delhi slums | |
| 46 | SP45 | Mapping Exercise of Kangaroo Mother Care | YES | YES | C | NO | | NO | | NO | | Focus on training | |
| 47 | SP46 | A KMC E-learning Platform as an effective t | YES | YES | C | NO | | NO | | NO | | Focus on e-learning platform and data platform [R | x |
| 48 | SP47 | KANGAROO CARE COVERAGE: FROM PILOT PI | YES | YES | C | NO | | NO | | NO | | Focus on training and expansion of tutors and hos | x |
| 49 | SP48 | UGANDA: Establishing a Regional Learning | NO | NO | E | NO | | NO | | NO | | Description of learning network - how it is done - n | x |
| 50 | SP49 | MALAWI: Using Program-Based Evidence ai | YES | | C | NO | | NO | | NO | | Focus on mentoring | |
| 51 | SP50 | Ensuring Quality of Care for Small Babies in | YES | YES | C | YES | p 14-16 perc | NO | | NO | | Comprehensive approach | |
| 52 | SP51 | Assessing and improving quality of care for | NO | NO | E | NO | | NO | | NO | | Comprehensive approach p32 "KMC should not be | x |
| 53 | SP52 | KMC Implementation Experience MCSP –Ta | NO | NO | E | YES | p8 Admissio | NO | | NO | pp 9-10 Conf | x | |
| 54 | SP53a& | Implementing Kangaroo Mother Care (KMC | INDIRECTLY | NO | C | NO | | NO | | NO | | Presentation and a more comprehensive text | |
| 55 | SP54 | Health Facility Assessment for Care of Prer | NO | NO | C | YES | p4 14% of eli | NO | | NO | | Criteria for facility readiness - see also AS31 | |
| 56 | SP55 | Summary of the conclusions of the worksh | NO | NO | E | NO | | NO | | NO | | Workshop topics | |
| 57 | SP56 | Kangaroo mother care Sustainability and f | INDIRECTLY | YES | C | NO | | NO | | NO | | | |
| 58 | SP57 | THE FUTURE OF KMC: ROLE OF NGO's The Ph | INDIRECTLY | YES | E | NO | | NO | | NO | | Role of NGO: focus on advocacy, training and othe | x |
| 59 | SP58 | ROLE OF NGOS IN PROMOTION OF KMC EXPE | INDIRECTLY | YES | E | NO | | NO | | NO | | Role of NGO: focus on promotion and training | |
| 60 | SP59 | IMPROVING AVAILABILITY, QUALITY AND USE | YES | YES | B | YES | p9 "Estimate | NO | | NO | | HMIS - check with R43 | |
| 61 | SP60 | Use of routine data: Monitoring KMC throu | NO | NO | D | YES | p19 "ESTIMAT | YES | p24 ">80% of | YES | p24 | Also check notes at the bottom of each slide | |
| 62 | SP61 | Estrategia Programas Madre Canguro en C | YES | YES | E | | | NO | | NO | | | |
| 63 | SP62 | Lineamientos técnicos para la implement | NO | NO | E | | | NO | | NO | | | |
| 64 | | | | | | | | | | | | | |
| 65 | 33 | | | | | | | | | | | | |
| 66 | 53.2% | Independent author/s | | | ↓ A,B,C,D,E,Missing | | | | | | | | |
| 67 | | Media House | 25 | 24 | 0 | 5 | YES | 1 | | 1 | | | |
| 68 | | Funding agency | 30 | 37 | 2 | 55 | NO | 61 | | 61 | | | |
| 69 | | Development agency | 0 | 0 | 25 | 0 | UNSURE | 0 | | 0 | | | |
| 70 | | Govt | 0 | 0 | 8 | 0 | N/A | 0 | | 0 | | | |
| 71 | | Other | 7 | 0 | 27 | 0 | INDIRECTLY | 0 | | 0 | | | |
| 72 | | Missing | 0 | 1 | 0 | 2 | Missing | 0 | | 0 | | | |
| 73 | | Total | 62 | 62 | 62 | 62 | Total | 62 | | 62 | | | |

(AB) Abstracts (49) | (AS) Assessments (31) | (R) Reports (56) | **(SP) Slides & pres (62)** | (SC) Scal ...

#### (SC) Scale-up (and implementation guides)

| | A | B | C | D | E | F | G | H | I | J | K |
|---|---|---|---|---|---|---|---|---|---|---|---|
| 1 | | 2 Title | Author | Publisher | Place of publication | Date published | URL | URL date accessed | Types of document | REMARKS | 17 Anal compl NVIVO (Date) |
| 2 | SC01 | Kangaroo Mother Care Implementation G | USAID/MCHIP/ Save | MCHIP Jhpiego | Washington DC | 2012 | https://www.mchip. | 2021/09/09 | Guide | | 2021/09/09 |
| 3 | SC02 | Implementation Workbook for Kangaroo I | Anne-Marie Bergh | MRC Unit for Materr | Pretoria, South Afri | 2002 | https://www.health | 2021/09/19 | Guide | | 2021/09/19 |
| 4 | SC03 | Accelerating the effective scaling of Kang | Nathaniel Foote, G | Early Childhood Ma | | 2017 | https://bernardvanl | 2021/08/10 | Article | | 2021/09/11 |
| 5 | SC04 | KANGAROO MOTHER CARE & OPTIMAL FEEI | GOI | Child Health Divisic | New Delhi | September 2014 | https://nhm.gov.in/ | 2021/09/15 | Guide | 2 columns | 2021/09/19 |
| 6 | SC05 | KANGAROO MOTHER CARE: Clinical Implen | Kenya Ministry of H | Kenya Ministry of H | (Nairobi) | 2016 | - | - | Guide | | 2021/09/13 |
| 7 | SC06 | Atenção Humanizada ao Recém-Nascido | Ministry of Health | MINISTRY OF HEALTI | Brasilia | 2017 (3rd ed) | https://bvsms.saud | 2021/09/19 | Guide | Module 1 English translation: 1 The Kangaroo M | 2021/09/19 |
| 8 | SC07 | Introducing and sustaining EENC in hospi | WHO WPR | World Health Orgar | Manila | 2018 | https://apps.who.in | 2021/09/19 | Guide | First Embrace; tables | 2021/09/20 |
| 9 | SC08 | THE PATHWAY TO HIGH EFFECTIVE COVERA | Lara Vaz, Tanya Gu | Save the Children, E | | no date | https://resourcecen | 2021/09/20 | Poster | NB for scale up ideas; poster; 3 columns | 2021/09/20 |
| 10 | SC09a | Scaling up kangaroo mother care in the P | Anthony Patrick Cal | BMJ Global Health | | 2021 | https://gh.bmj.com/ | 2021/09/20 | Article | 2 columns; BMJ Global Health 2021;6:e006492. dc | 2021/09/20 |
| 11 | SC09b | Capacity Development on Hospital Policy | Presenters listed | | | no date | - | - | Other | Received from Sookee | 2021/09/20 |
| 12 | SC10 | Lessons learned from the introduction ar | Joseph de Graft-Jol | Save the Children, A | | 2007 | https://www.health | 2021/09/20 | Poster | Poster; 3 columns | 2021/09/20 |
| 13 | SC11 | KMC Acceleration Framework | Save the Children | | | June 2015 | https://www.health | 2021/09/20 | Other | NB Only a diagram; includes the buiding blocks t | 2021/09/20 |
| 14 | SC12 | Kangaroo Mother Care—High on Evidence | Amrita Misra, Sarat | JSI | | 12 Oct 2017 | https://www.jsi.com | 2021/09/20 | Brief | Viewpoint (brief) | 2021/09/20 |
| 15 | SC13 | JSI CONTRIBUTES TO SCALE-UP OF KANGARO | JSI | | | no date | https://www.jsi.com | 2021/09/20 | Infographic | Infographic | 2021/09/20 |
| 16 | SC14 | Informing the design of a trial of kangarc | Dr Melissa Morgan | LSHTM | London | August 2020 | https://researchonl | 2021/09/20 | Thesis | PhD thesis | 2021/09/20 |
| 17 | SC15 | 'Scaling-up is a craft not a science': Catal | Spicer N, Bhattacha | Soc Sci Med | | 2014 | https://www.scienc | 2021/09/22 | Article | 2 columns; Soc Sc Med 2014;121:30-38; KMC not m | 2021/09/22 |
| 18 | SC16 | National Newborn Health Program Imple | Ministry of Health | MoHFW | Bangladesh | 2017 (1st ed); 2019 | https://www.resear | 2021/09/22 | Guide | Extracts taken | 2021/09/23 |
| 19 | SC17a | Scaling up Kangaroo Mother Care in Ethio | Mony PK, Tadele H, | BMJ Global Health | | 2021 | https://gh.bmj.com/ | 2021/09/22 | Article | 2 columns; BMJ Global Health. 2021; 6: e005905 | 2021/09/22 |
| 20 | SC17b | Scaling up Kangaroo Mother Care - supple | Mony PK, Tadele H, | BMJ Global Health | | 2021 | https://gh.bmj.com/ | 2021/09/22 | Supplement | BMJ Global Health. 2021; 6: e005905 | 2021/09/23 |
| 21 | | | | | | | | | | | |
| 22 | 19 | | | Journal publications | | | | Guide | 7 | | |
| 23 | 100% | | | | | | | Article | 4 | | |
| 24 | | | | | | | | Supplement to article | 1 | | |
| 25 | | | | | | | | Poster | 2 | | |
| 26 | | | | | | | | Thesis | 1 | | |
| 27 | | | | | | | | Brief | 1 | | |
| 28 | | | | | | | | Infogprahic | 1 | | |
| 29 | | | | | | | | Other | 2 | | |
| 30 | | | | | | | | | 19 | | |
| 31 | | | | | | | | | | | |
| 32 | | | | | | | | | | | |
| 33 | | | | | | | | | | | |
| 34 | | | | | | | | | | | |
| 35 | | | | | | | | | | | |
| 36 | | | | | | | | | | | |

... (AS) Assessments (31) | (R) Reports (53) | (SP) Slides & pres (60) | **(SC) Scale-up & impl (19)** | TE ...
