## Supplemental file 4 for "Programmatic implementation of kangaroo mother care: a systematic synthesis of grey literature"

Supplemental file 4 Documents analysed with NVivo

| | | File name | Nodes | References |
|---|---|---|---|---|
| | 1 | AS01 HNN 10-Nigeria-2011-KMC-review-report_0 2021-08-07 | 31 | 136 |
| | 2 | AS06 MCSP Dominican Republic -KMCAssessmentReport - 2021-07-23 | 30 | 108 |
| | 3 | AS07 MCSP Myanmar_Feasibility & Acceptability of KMC at Taunggyi_Report 2021-07-27 | 31 | 126 |
| | 4 | AS08 MCSP Malawi-BNA-Report-FINAL 2021-07-23 | 32 | 174 |
| | 5 | AS10 UNICEF SURVEY 25 countries - long manuscript with tables & boxes | 18 | 32 |
| | 6 | AS11 South Africa NW Prov KMC survey 2015 presentation | 25 | 52 |
| | 7 | AS12 UNICEF Chad Trip report FINAL | 28 | 49 |
| | 8 | AS13 UNICEF Togo Trip report ENGLISH Final draft | 31 | 47 |
| | 9 | AS14 Indonesia 2009-10 KMC FINAL REPORT - Final_Sep28 - confidential (AMB) | 33 | 86 |
| | 10 | AS15 SC Malawi KMC Retrospective Study Report 2008 - fInal-final | 26 | 47 |
| | 11 | AS16 SNL Brief Tracking Implementation Progress for KMC 2021-07-23 | 25 | 52 |
| | 12 | AS16a SNL evaluation 2012 Multi-country KMC evaluation brief v2 | 8 | 9 |
| | 13 | AS17 SNL evaluation 2012 Malawi KMC report - FINAL FINAL | 14 | 16 |
| | 14 | AS18 SNL evaluation 2012 Mali KMC Report | 19 | 37 |
| | 15 | AS19 SNL evaluation 2012 Rwanda KMC report - FINAL 1 | 22 | 39 |
| | 16 | AS20 SNL evaluation 2012 Uganda KMC report - FINAL | 19 | 26 |
| | 17 | AS21 Deep Dive 2015-01-05 KMC Asia consolidated report FINAL -table disabled | 46 | 160 |
| | 18 | AS22 Deep dive Asia India KMC report FINAL 2015-04-15 | 9 | 13 |
| | 19 | AS23 Deep Dive Asia KMC Assessment Philippines_Final Report 5 | 35 | 135 |
| | | AS24 Deep Dive Asia kmc indonesia final 20 jan hp - CANNOT OPEN | 0 | 0 |
| | 20 | AS25 Deep Dive Asia KMC Pakistan report - FINAL - 2014-11-12 | 20 | 88 |
| | 21 | AS26 Deep Dive Asia Situation Analysis_KMC_Bangladesh_April 24 2014_the final | 0 | 0 |
| | | AS27 SC Nigeria KMC-brief-18Aug17-A4-LOGOS (1) CANNOT PROCESS | 0 | 0 |
| | | AS27a SC Nigeria KMC-brief-18Aug17-A4-LOGOS (1) CANNOT PROCESS | 0 | 0 |
| | 22 | AS28 SC Nigeria Report Care of Small Newborns final (2017)+C53 | 49 | 398 |
| | 23 | AS31 HNN Ehtiopia KMC_Facilities_Brief_ 2021-07-24 | 17 | 26 |
| | 24 | SC01 MCHIP KMC Implementation Guide 2021 | 31 | 120 |
| | 25 | SC02a C KMC WORKBOOK - CONTENTS, INTRO - REFORMATTED | 4 | 7 |
| | | SC02b D KMC WORKBOOK - PART 1 - REFORMATTED | 14 | 20 |
| | | SC02c E KMC WORKBOOK- PARTS 2 & 3 - REFORMATTED | 17 | 28 |
| | | SC02d F KMC WORKBOOK - PART 4 - REFORMATTED | 19 | 28 |
| | | SC02e G KMC WORKBOOK - PART 5 - REFORMATTED | 7 | 12 |
| | | SC02f H KMC WORKBOOK - PARTS 6 & 7 - REFORMATTED | 9 | 16 |
| | | SC02g I KMC WORKBOOK - HANDOUTS - REFORMATTED | 1 | 1 |
| | 26 | SC03 ECM17_11_KMC_Foote | 11 | 19 |
| | 27 | SC04 (G1) KENYA KANGAROO+MOTHER+CARE+IMPLEMENTATION+GUIDELINES+2016(1) (002) | 19 | 66 |
| | 28 | SC05 (G2) India Operational_Guidelines-KMC_&_Optimal_feeding_of_Low_Birth_Weight_Infants 2021-07-20 | 28 | 145 |
| | 29 | SC06 Brazil Atencao_humanizada_metodo_canguru_MODULE 1 ENGLISH 2021-07-31 | 10 | 29 |
| | 30 | SC07 HNN First Embrace Introducing and sustaining EENC in hospitals - teaching module 2021-07-24 | 23 | 69 |
| | 31 | SC08 SC Pathway_to scale up poster 2021-07-30 | 8 | 26 |
| | 32 | SC09a Calibo Philippines KMC scale up BMJ Glob Health 2021 | 25 | 79 |
| | 33 | SC09b Philippines 6 CSBPOLICY_Schedule of Activities_revised (1) | 7 | 11 |
| | 34 | SC10 HNN Lessons KMC expansion poster_final 2021-07-25 | 13 | 18 |
| | 35 | SC11 HNN SC KMC Acceleration Framework 2021-07-24 | 10 | 17 |
| | 36 | SC12 JSI India Viewpoint Kangaroo Mother Care–High on Evidence, Low on Reach | 10 | 13 |
| | 37 | SC13 JSI India Vriddhi RMNCH-A KMC-Infographic 2021-07-24 | 2 | 2 |
| | 38 | SC14 Medvevdev-2020_Informing_the_design_of_a_trial - scale-up | 31 | 116 |
| | 39 | SC15 NB HNN Scaling Up Craft Not Science_SocSciMedicine | 22 | 87 |
| | 40 | SC16a Bangladesh NNHP Implementation Toolkit_English EXTRACTS | 20 | 51 |
| | 41 | SC17a Mony Scaling_up_KMC_Ethiopia_and_India BMJ GH 2021 | 25 | 71 |
| | 42 | SC17b Mony bmjgh-2021-005905supp001_data_supplement | 10 | 24 |

37 Grey documents
4 Articles
1 Article supplement
