## Supplemental file 5 for "Programmatic implementation of kangaroo mother care: a systematic synthesis of grey literature"

**NODES CREATED ON NVIVO**

Background & problem *See also KMC & global health*

Bangladesh

Barriers – obstacles [including challenges; weaknesses; bottlenecks] *See also Situation analysis; Implementation conditions for*

Benchmarking

Beyond KMC – ECD [including first 1000 days]

Capacity building [including training general; orientation; clinical coaching; pre-service & in-service; curriculum/a; mentorship; competencies; skills; knowledge; knowledge and skills dissemination; TOT] *See also Supervision; Health workforce; Training materials*

Communication *See also Integration*

Community [including demand side; civil society; community health workers (CHWs); fathers; male involvement; mobilisation; access; engagement] *See also IEC (materials)*

Context [including complexity; no one-size-fits-all; dependency; health system issues - general]

Country names [including more detail on Myanmar; Chad; Togo; Madagascar; South Africa; Iran; Pakistan]

Coverage [including uptake; patronage] [sub-node: lack of coverage] *See also Service provision*

Cultural & personal issues [including acceptability for parents and community; (socio)cultural barriers; adherence; compliance; resistance to KMC] *See also IEC (materials)*

Dominican Republic

Education (mothers; parents; family) [including family counselling; psychosocial support; all job aids] *See also Service provision; IEC materials; Support family*

Equipment and supplies [including food; meals]

Ethiopia

Evidence

Facilitators [including enablers; supporting factors] *See also Implementation conditions for; Situation analysis*

Family participation *See also Support family*

Feeding and growth (KMC nutrition) [including breastfeeding; growth monitoring; BFHI; weight …]

Finances [including external / donor funding; investment; accreditation / certification; national health insurance; costing; budget; donors] *See also Resources*

Follow-up [including (neurological / motor development and ROP) evaluations] *See also Referral*

Foundations and networks

Guidelines & policies [including strategic plans; ordinances; regulations; decrees (health system or facility level); acts; protocols; SOPs; ENAP] *See also Implementation clinical and practical; Education (job aids)*

History KMC

Health workforce [including health workforce; support for staff; rotations; acceptability by staff; resistance] *See also Capacity building; Role players*

IEC (materials) [including messages; international days; use of celebrities; social and behaviour communication and change (SBCC); (mass) media] *See also Education; Community; Cultural and personal issues*

Implementation clinical & practical [including position and discharge; admission; types of KMC; adherence; handling of small newborns] *See also Guidelines and protocols; Thermal care (non-KMC)*

Implementation conditions for [including key actions; conditions / factors for scale-up; prerequisites; attributes; catalyse; certification / accreditation; barriers & facilitators sides of the same coin; determinants of KMC implementation]

Implementation pathways [including roadmaps] *See also Models implementation & scale-up*

Implementation progress *See also Monitoring and evaluation*

India

Indonesia

Infrastructure [including space, lodger mothers]

Integration [including continuum / continuity of care; linkages; continuation across points of care; harmonisation; collaboration; nurturing care] *See also Communication*

KMC & global health *See also Background & problem*

Leadership & governance [including partnership issues; political will; decentralisation; health system organisation; management; accountability] *See also Integration (harmonisation)*

Levels of care

Malawi

Miscellaneous [including innovations; (MNH) interventions – general; broader societal issues]

Models implementation & scale-up [including paradigm shifts; approaches; centre of excellence; model sites; roadmaps] *See also Scale-up preparation; Plan of action*

Monitoring and evaluation [including health information systems; HMIS; using data for improving services indicators; accountability] *See also Records; Implementation progress*

Nepal

Nigeria

Philippines

Plan of action [sub-node: Plan] [including specific topics/areas to be attended to] *See also Models implementation & scale-up*

Prioritisation

Quality of care – improvement [including certification] *See also Monitoring and evaluation*

Records [including registers; forms; accountability] *See also Monitoring and evaluation*

Referral *See also Transport; Follow-up*

Resources *See also Finances*

Role players [including multidisciplinary team(work); managers; facilitators; partners(hips); champions; implementers; driver; focal person funders; donors; professionals; private hospitals; engagement; collaboration] *See also Health workforce; Stakeholders*

Scale-up description-definition

Scale-up general

Scale-up preparation [including planning; next steps; roadmap] *See also Plan of action; Models implementation & scale-up*

Service provision [including service delivery; organisation of services; (KMC) programming; coverage / availability of (KMC) services; hygiene / WASH] *See also Education (job aids); Coverage; Support family*

Situation analysis – assessment [including research / link implementation with research; readiness at all levels; bottleneck analysis; baseline] *See also Barriers – obstacles; Facilitators*

South Africa

Stage 1 Awareness [including advocacy; sensitisation] *See also IEC (materials)*

Stage 2 Commitment

Stage 3 *See Scale-up planning*

Stages of change

Stakeholders *See also Role players*

Supervision [including supportive supervision] *See also Capacity building*

Support family [including guardians; food; enabling families to do KMC] *See also Education; Service provision; Family participation*

Thermal care (non-KMC) *See also Implementation clinical & practical*

Tools

Training Materials [including educational materials] *See also Capacity building*

Transport *See also Referral*

What is KMC?
