## Supplemental file 6 for "Programmatic implementation of kangaroo mother care: a systematic synthesis of grey literature"

Supplemental files 6 Document types and levels of importance

**Documents types for first round analysis**

| Document type | Abstracts (posters, summaries, working groups) | | Assessments, evaluations and reviews | | Reports, briefs, newsletters & blogs | | Slides & conference presentations | | Implementation and scale-up documents | | TOTAL | |
|---|---|---|---|---|---|---|---|---|---|---|---|---|
| | n | % | n | % | n | % | n | % | n | % | n | % |
| Report | 19 | 39% | 27 | 87% | 25 | 45% | 0 | 0% | 0 | 0% | 71 | 33% |
| Brief | 0 | 0% | 4 | 13% | 16 | 29% | 0 | 0% | 1 | 5% | 21 | 10% |
| Conference abstract | 23 | 47% | 0 | 0% | 0 | 0% | 0 | 0% | 0 | 0% | 23 | 11% |
| Poster | 7 | 14% | 0 | 0% | 0 | 0% | 0 | 0% | 2 | 11% | 9 | 4% |
| Conference presentation | 0 | 0% | 0 | 0% | 0 | 0% | 62 | 100% | 0 | 0% | 62 | 29% |
| Blog | 0 | 0% | 0 | 0% | 3 | 5% | 0 | 0% | 0 | 0% | 3 | 1% |
| Newsletter | 0 | 0% | 0 | 0% | 7 | 13% | 0 | 0% | 0 | 0% | 7 | 3% |
| Dissertation/thesis | 0 | 0% | 0 | 0% | 1 | 2% | 0 | 0% | 1 | 5% | 2 | 1% |
| Other | 0 | 0% | 0 | 0% | 4 | 7% | 0 | 0% | 15 | 79% | 19 | 9% |
| **TOTAL** | **49** | **100%** | **31** | **100%** | **56** | **100%** | **62** | **100%** | **19** | **100%** | **217** | **100%** |

**Documents analysed in the first round**

| | | In-depth | | Nuggets | | Nothing | |
|---|---|---|---|---|---|---|---|
| (AB) Abstracts (also posters, summaries & working groups) | **49** | 16 | 32.7% | 17 | 34.7% | 16 | 32.7% |
| (AS) Assessments, evaluations and reviews | **31** | 24 | 77.4% | 7 | 22.6% | 0 | 0.0% |
| (R) Reports, briefs, newsletters & blogs | **56** | 27 | 48.2% | 24 | 42.9% | 5 | 8.9% |
| (SP) Slides & conference presentations | **62** | 32 | 51.6% | 14 | 22.6% | 16 | 25.8% |
| (SC) Scale-up (and implementation guides) | **19** | 19 | 100.0% | 0 | 0.0% | 0 | 0.0% |
| **TOTAL** | **217** | **118** | | **62** | | **37** | |
